# Towards the Virtual Amyotrophic Lateral Sclerosis Patient: Inferring Cortical Excitability through Whole-Brain Dynamical Modeling

**DOI:** 10.64898/2026.06.09.26354829

**Authors:** Marianna Angiolelli, Matteo Demuru, Emahnuel Troisi Lopez, Meysam Hashemi, Abolfazl Ziaeemehr, Giovanni Rabuffo, Francesca Trojsi, Carmine Granata, Domenico Tafuri, Mario De Luca, Enrica Gallo, Giuseppe Sorrentino, Viktor Jirsa, Damien Depannemaecker, Pierpaolo Sorrentino

## Abstract

Amyotrophic Lateral Sclerosis (ALS) is increasingly recognized as a multisystem neurodegenerative disorder in which motor-neuron degeneration is accompanied by widespread alterations in cortical dynamics. Hyperexcitability is widespread across the brain, although more predominantly in motor and premotor areas. Yet, how local excitability imbalance shapes distributed whole-brain dynamics remains poorly understood. Here, we combined source-reconstructed resting-state magnetoencephalography data, tractography-informed whole-brain modeling, and simulation-based inference to investigate whether ALS-related alterations in large-scale brain dynamics can be mechanistically explained by changes in cortical excitability.

First, we characterized empirical brain dynamics using neuronal avalanche transition matrices (ATMs) on source-reconstructed magnetoencephalography data. These analyses revealed significant alterations in ALS patients relative to healthy controls. To interpret these changes mechanistically, we employed a reduced Wong–Wang whole-brain model in which local recurrent excitation modulates emergent large-scale neural dynamics. The model showed that increasing recurrent excitation uniformly across the brain produced changes in avalanche-derived features that were directionally consistent with the empirical alterations observed in ALS patients. Then, using simulation-based inference, we jointly estimated recurrent excitation and global coupling from individual empirical feature vectors. ALS patients exhibited higher inferred recurrent excitation than controls. A regional extension of the model, distinguishing motor and frontal from non-frontal regions, revealed more pronounced hyperexcitability in motor-frontal regions. Together, these findings support enhanced excitability as a candidate mechanism contributing to altered large-scale avalanche propagation in ALS. By integrating empirical brain dynamics with probabilistic model inversion, this framework provides individual-level mechanistic descriptors of disease-related brain activity and motivates further investigation of regional excitability alterations and their clinical relevance.

## 1 Introduction

Amyotrophic Lateral Sclerosis (ALS) is a progressive neurodegenerative disorder characterized by the degeneration of both upper and lower motor neurons, ultimately leading to muscle weakness, atrophy, and death [1]. Although its clinical manifestations are well described, the underlying pathophysiological mechanisms remain only partially understood.

One of the most consistent neurophysiological findings in ALS is cortical hyperexcitability [2–4], with potential utility for prognosis and stratification [5]. Transcranial magnetic stimulation (TMS) can be used to assess short-interval intracortical inhibition (SICI), which indexes cortical excitability, by probing GABA-A-mediated inhibitory interneuronal circuitry [6–8]. Reduced SICI reflects diminished inhibitory control and the consequent increase in neuronal recruitment. SICI is diminished in ALS, both idiopathic and familial [9–11], before lower motor neuron impairment [2]. Increased cortical hyperexcitability has been linked to greater functional impairment, as measured by ALSFRS-R scores [12], while reduced average SICI independently predicts shorter survival in early-stage patients [13–15].

Although cortical hyperexcitability has been extensively investigated in motor circuits, ALS-related alterations also involve regions beyond the motor system [16–19]. Accordingly, the reorganization of brain dynamics extends beyond motor areas, encompassing the entire brain [20], and is strongly associated with clinical impairment [20] and with disease staging [21]. These observations motivate investigation of how local circuit alterations interact with distributed network organization to shape whole-brain dysfunction. This points to widespread changes in meso-scale circuitry that reverberate across the whole brain. Together, these observations support the hypothesis that whole brain dysfunction in ALS is best understood as the result of the interaction between local cortical hyperexcitability and the active, potentially compensatory or maladaptive, functional reorganization of distributed brain networks.

However, the link between local cortical hyperexcitability and distributed brain dysfunction remains difficult to establish from empirical measurements alone. TMS-derived metrics provide a powerful and clinically relevant index of inhibitory dysfunction within motor cortical circuits, but they do not directly reveal how altered excitation-inhibition reshapes whole-brain dynamics. Conversely, whole-brain electrophysiological or imaging markers can capture distributed reorganization, but they remain largely descriptive unless embedded within a framework that explicitly models the underlying biophysical mechanisms.

These limitations motivate the adoption of model-based approaches [22]. In fact, mechanistic whole-brain models allow us to move beyond purely correlational analyses by simulating how different, non-directly observable neurodegenerative processes may give rise to measurable large-scale brain dynamics [23–25]. By comparing these simulated dynamics, generated by “virtual brain twins” with data recorded from individual patients, one can estimate latent pathophysiological variables, thereby characterizing the pathophysiological processes underlying the observed brain activity.

We hypothesized that model inversion would identify a recurrent-excitation parameter associated with ALS-related alterations in resting-state brain dynamics. We further hypothesize that the correlation between motor-frontal hyperexcitability and clinically relevant features would be more prominent than that with non-frontal regions. In this study, we analyzed a cohort of 39 patients diagnosed with ALS and 39 controls, for whom clinical parameters, source-reconstructed magnetoencephalography, and magnetic resonance data were acquired, as in [20]. First, we assessed whether features extracted from resting-state brain activity could distinguish ALS patients from controls and whether these alterations were associated with clinical outcomes. The characterization of clinical features was performed under the hypothesis that aperiodic bursts of activity (i.e., neuronal avalanches) would index pathophysiological processess [20, 26] . This empirical characterization provided the basis for subsequent computational modeling and model-inversion analyses. To test whether the observed changes in local excitability could explain the observed shifts in large-scale brain dynamics, we used the reduced Wong–Wang model [27,28], which describes the mean synaptic activity of interacting excitatory populations. Whole-brain neural activity was simulated by coupling neural masses via structural connectomes derived from diffusion-weighted tractography [29, 30]. Because of its low-dimensional formulation, this model is particularly well-suited to whole-brain simulations aimed at linking regional excitability-related parameters to emergent macroscale neural dynamics. Along these lines, rather than explicitly introducing a separate parameter for inhibition, we abstracted the local excitation–inhibition balance within each cortical region through the recurrent excitation parameter. Two complementary model configurations were examined. In the first, we assumed spatially uniform recurrent excitation, and jointly inferred the global recurrent-excitation parameter *w* and the global coupling parameter *G*. This configuration allowed us to test whether ALS-related alterations in whole-brain dynamics were compatible with an overall increase [31] in recurrent excitation while accounting for the contribution of interregional coupling. In the second model configuration, we divided the brain into two anatomically defined partitions: a motor-frontal partition, including frontal and motor cortical areas, and a complementary partition comprising the remaining regions. The second model configuration was motivated by the heterogeneous anatomical distribution of ALS-related pathology. Although ALS primarily affects the motor system, including the primary motor cortex and corticospinal pathways, premotor and frontal association regions are also frequently involved, with the latter particularly implicated in patients exhibiting cognitive and behavioral impairment [**?**, 32–35]. Conversely, although alterations may extend throughout the brain, the clinical manifestations of ALS are generally dominated by motor and, to a variable extent, frontal-executive dysfunction [36–38]. Based on these observations, we introduced a parsimonious anatomical subdivision into a motor-frontal partition comprising 34 regions, including primary motor, premotor, and frontal association cortices, and a complementary non-frontal partition comprising the remaining 46 regions. Importantly, this subdivision does not imply that regions outside the motor-frontal partition are unaffected, nor that all regions within it are equally involved. Rather, it provides a simplified, biologically motivated framework to test whether ALS-related alterations in local recurrent excitation are more pronounced in anterior regions than in the rest of the cortex. In particular, we hypothesized the motor-frontal regions to be more strongly hyperexcitable and proportional to individual clinical features. To test this hypothesis, separate recurrent-excitation parameters, *w_F_* and *w_NF_* , were assigned to the two partitions and jointly inferred with the global coupling parameter G. This allowed us to investigate the spatial heterogeneity of inferred recurrent excitation while accounting for interregional coupling. Both configurations were inferred independently using summary features extracted from whole-brain avalanche transition matrices. To this end, we adopted a Bayesian framework based on simulation-based inference (SBI) [39], which enables efficient inversion of complex generative models [40, 41]. This approach was applied at the individual level, allowing us to investigate both group-level alterations and interindividual variability in inferred parameters. Finally, we examined whether the estimated parameters were associated with clinical measures of functional impairment and disease staging. An overview of the computational framework is provided in Fig. 1.

**Figure 1:**
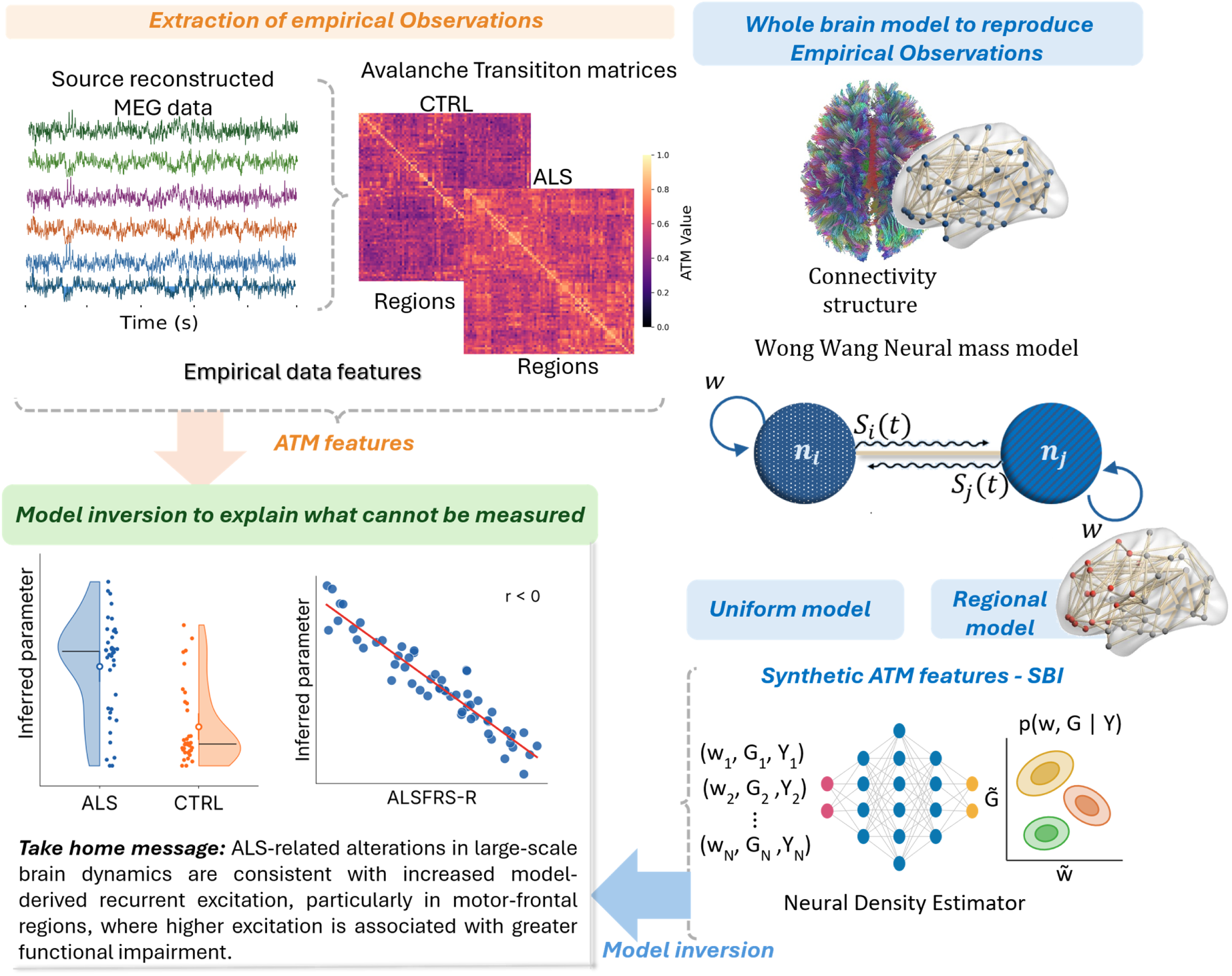
Overview of the computational framework for estimating latent cortical excitability in ALS. The framework integrates resting-state magnetoencephalography (MEG), connectome-based whole-brain modeling, and simulation-based inference (SBI) to link alterations in large-scale brain dynamics to underlying changes in recurrent cortical excitation. **Extraction of empirical observations.** Source-reconstructed resting-state MEG recordings from patients with amyotrophic lateral sclerosis (ALS) and healthy controls (CTRL) were used to characterize large-scale brain dynamics. Neuronal avalanches were identified from regional activity, and Avalanche Transition Matrices (ATMs) were constructed to quantify the propagation of activities across brain regions. Summary statistics derived from the empirical recordings were used as targets for model inversion. Six ATM-derived summary features were used for model inversion: the mean and median of the strictly upper and lower triangular entries, the Frobenius norm, and singular-value-based entropy. These features were extracted using the same procedures from empirical recordings and simulated activity as described in the Methods section, not shown here. **Whole-brain modeling and simulation-based inference.** A structural-connectivity-constrained reduced Wong-Wang neural mass model was used to generate synthetic brain activity across different combinations of model parameters. Two model configurations were investigated: a uniform model, characterized by a single global recurrent-excitation parameter, and a regional model, distinguishing recurrent excitation in motor-frontal and non-frontal regions. Synthetic signals were processed to extract the corresponding data features, which were used to train neural density estimators and approximate the posterior distributions of the model parameters conditioned on the empirical observations. **Model inversion and physiological interpretation.** Subject-specific posterior distributions were estimated by conditioning the trained inference framework on empirical features. The inferred parameters revealed increased model-derived recurrent excitation in ALS, particularly in motor-frontal regions, where higher excitation was associated with greater functional impairment. These findings suggest that ALS-related alterations in large-scale brain dynamics are consistent with changes in latent cortical excitation that macroscopic recordings cannot directly measure. **Abbreviations:** ALS, amyotrophic lateral sclerosis; CTRL, healthy controls; MEG, magnetoencephalography; ATM, Avalanche Transition Matrix; SBI, simulation-based inference; ALSFRS-R, Amyotrophic Lateral Sclerosis Functional Rating Scale–Revised.

## 2 Results

### 2.1 Characterization of large-scale dynamics of empirical data

In this study, we set out to identify informative features from MEG data acquired in ALS patients and controls (CTRL), and to produce a generative model to explain empirical data features as a function of local hyperexcitability. We identified neuronal avalanches as bursts of activities spreading across the whole brain, using a threshold-based approach [42], and turned to avalanche-based metrics (see also Methods). To characterize the spatial propagation of neuronal avalanches, we computed the Avalanche Transition Matrices (ATM) [43]. Each entry ATM*_ij_* encodes the probability that a suprathreshold activation in region *i* is followed by one in region *j* , within the same neuronal avalanche and then averaged across all avalanches. These matrices therefore describe how transient regional activations propagate through the brain. The ATM is intrinsically directed, as the probability that activation in region *i* is followed by activation in region *j* need not equal the probability of the reverse transition. Fig.2 displays the ATM averaged across ALS patients (Fig.2a) and healthy controls (Fig.2b). Warmer colors correspond to higher transition probabilities between brain regions. Fig.2 (c-h) report the distributions of the selected metrics of interest (MOIs). We computed the mean and median separately for the strictly upper and lower triangles, excluding the diagonal, and complemented these summaries with their Frobenius norm and entropy of the full matrix. Under the regional ordering held constant across participants and simulations, the two triangles summarize opposite directions for the same unordered pairs of regions. They provide complementary descriptions of the directed transition structure, rather than representing distinct anatomical subnetworks. Here, directionality refers to the temporal ordering of activations and does not establish causal influence between regions. The mean ATMs averages the transition probabilities across the regional pairs included in the analysis and therefore summarizes the overall propensity for activity propagation during avalanches. Higher mean ATM values indicate that suprathreshold activation in one region is, on average, more likely to be followed by suprathreshold activation in another region. ALS patients exhibited higher mean and median transition probabilities in the upper triangle than controls (Fig.2c, mean: Cohen’s *d* = 0.83, 95% CI [0.41, 1.29], *p* = 0.000523; panel e, median: *d* = 0.84, 95% CI [0.41, 1.32], *p* = 0.000480). In the lower triangle, the mean was also higher in ALS (Fig.2d) (*d* = 0.47, 95% CI [0.03, 0.95], *p* = 0.0419), whereas the median (Fig.2f) showed a smaller, nonsignificant difference in the same direction (*d* = 0.39, 95% CI [−0.05, 0.87], *p* = 0.0932). Together, these findings indicate higher average avalanche transition probabilities in ALS, consistent with an enhanced propensity for large-scale activity propagation. ATM entropy (Fig.2g) showed a negative but nonsignificant group difference (*d* = −0.27, 95% CI [−0.73, 0.18], *p* = 0.238). This measure quantifies the distribution of normalized singular values, with lower entropy indicating a greater concentration of spectral weight in fewer dominant components. Higher entropy reflects a more even contribution of multiple spectral modes. Stronger propagation does not necessarily imply greater spectral diversity, as transition probabilities may increase without substantially changing their relative spectral organization. Although the negative effect estimate was consistent with a greater concentration of spectral weight in dominant modes, the group difference was not statistically significant. Together, these findings indicate that group differences were more pronounced in the strength of avalanche propagation than in the spectral organization of the underlying transition matrix. The Frobenius norm of the ATM (Fig.2h) was higher in ALS patients (*d* = 0.55, 95% CI [0.12, 1.03], *p* = 0.0182) , indicating an increased overall magnitude of avalanche transition probabilities. The convergent increases in mean, median, and norm support an amplification of avalanche-mediated activity spreading at the whole-brain level, compatible with the hypothesis of cortical hyperexcitability in ALS. We additionally investigated whether the six empirical ATM features were associated with functional impairment and disease staging in ALS patients. Spearman rank-order correlations were computed between each feature and ALSFRS-R, King’s stage, and MiToS stage (Supplementary Fig. SM1). Although some associations were observed, only the median of the upper-triangular entries was associated with King’s stage at the nominal significance level (Spearman’s *ρ* = 0.345, unadjusted *p* = 0.0425, *q* = 0.1102). None of the associations survived Benjamini–Hochberg correction across the three clinical scales within each metric. Thus, the empirical ATM features revealed alterations in large-scale brain dynamics at the group level but did not exhibit detectable corrected associations with the examined clinical outcomes when considered individually.

**Figure 2:**
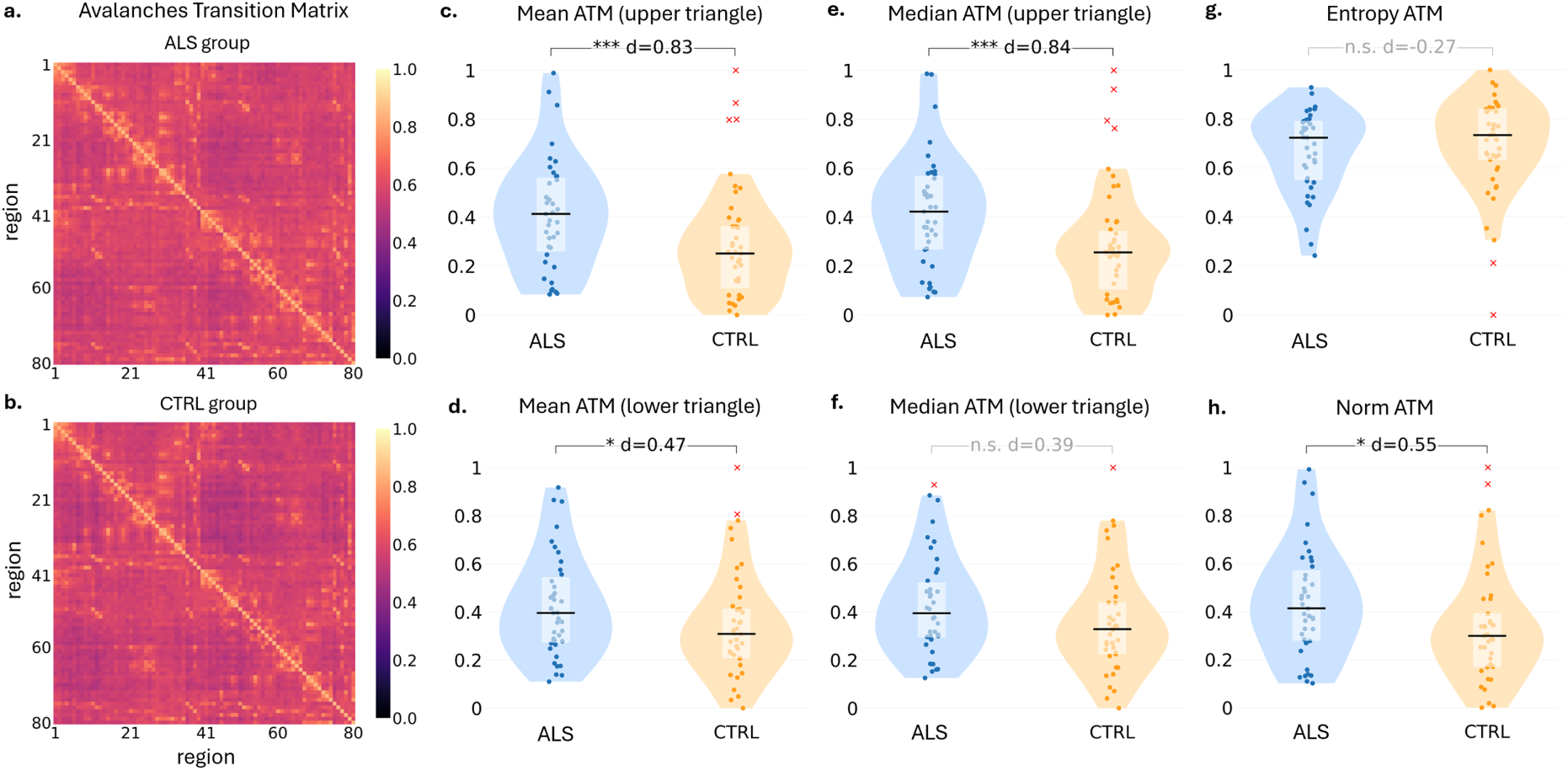
Whole-brain data features that characterize alterations in large-scale brain dynamics in ALS patients. Group-averaged Avalanche Transition Matrix (ATM) for ALS patients (**a**) and healthy controls (**b**). Outliers were excluded prior to averaging. Each matrix entry *ATM_ij_* represents the probability that a suprathreshold activation in region *i* is followed by an activation in region *j* during the same neuronal avalanche; warmer colors indicate higher transition probabilities. **(c–f)** Subject-level mean and median transition probabilities were computed separately from the strictly upper and lower triangles, excluding the diagonal. Mean values are shown in the left column and medians in the right column, with upper-triangle summaries above lower-triangle summaries. **(g,h)** SVD-based entropy and Frobenius norm of the full ATM, respectively. Violin plots show the distributions for ALS patients (blue) and controls (orange). Each dot represents one participant; white boxes indicate the interquartile range and black horizontal lines indicate the median. For visualization, each metric was independently min-max normalized to [0, 1] using the pooled ALS and control values, including outliers. Red crosses correspond to subjects identified as outliers using the interquartile range (IQR) criterion determined separately for each group and metric. These subjects were excluded from both the statistical group comparisons and the correlation analyses with clinical outcomes. Brackets report Cohen’s *d* and significance from two-sided Welch tests on the retained observations. Asterisks denote uncorrected *p*-values: n.s., *p* 0.05; * *p <* 0.05; ** *p <* 0.01; *** *p <* 0.001. Clinical correlation plots are reported in the Supplementary Material SM1.

### 2.2 Computational model and simulation-based inference to investigate a mechanistic substrate of altered ALS brain dynamics

To mechanistically interpret the empirical alterations described above, we investigated whether the large-scale dynamical signatures extracted from resting-state MEG recordings could emerge from changes in recurrent excitation and interregional coupling within a tractography-informed whole-brain model. We employed a reduced Wong-Wang neural mass model in which regional excitatory populations interact through empirical structural connectivity (Fig. 3a; see Methods).

**Figure 3:**
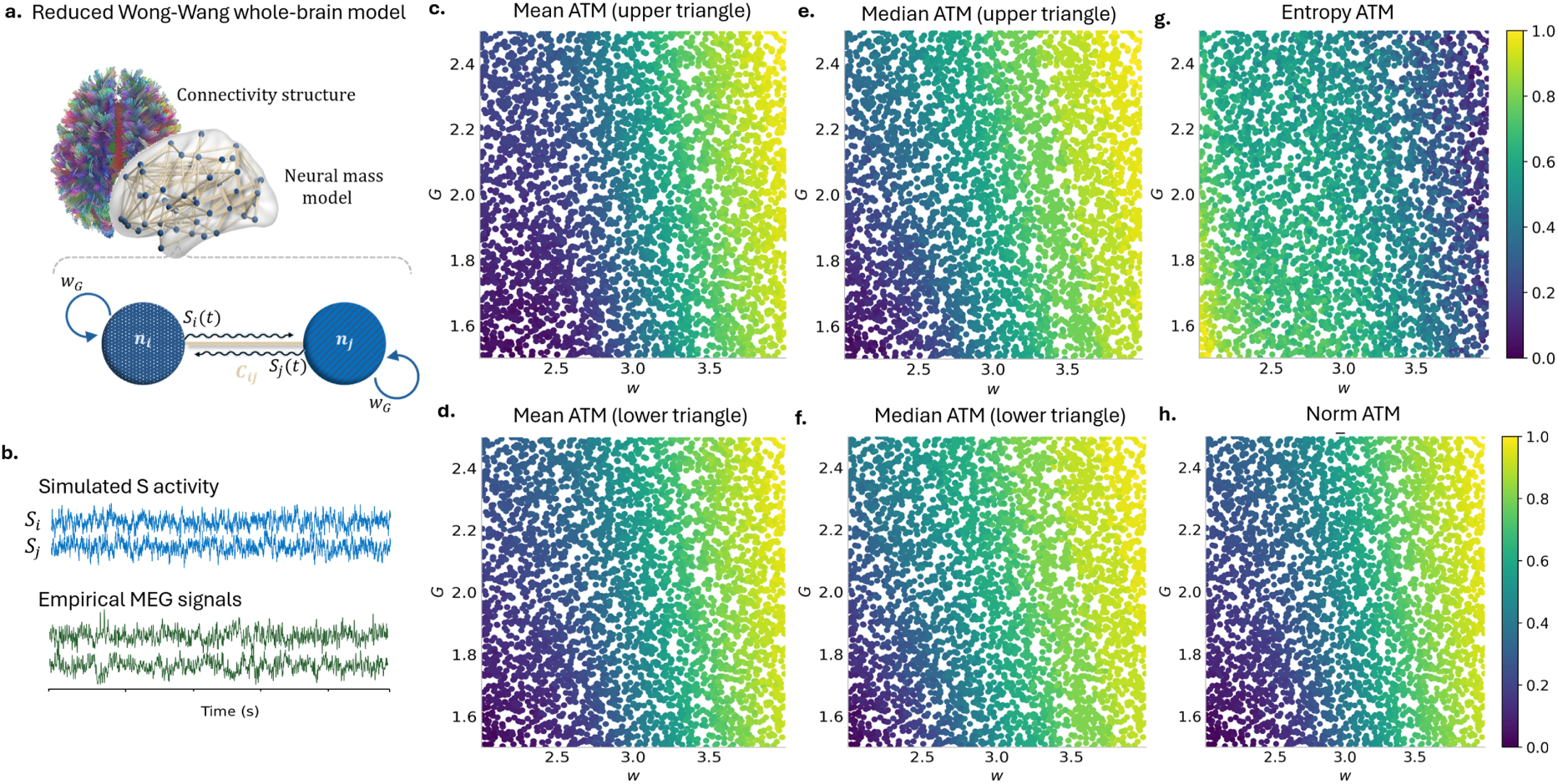
Tractography-informed whole-brain neural mass simulations link cortical excitability to ALS-specific dynamics. (**a**) Whole-brain neural activity was simulated using the Wong-Wang neural mass model informed by structural connectomes derived from diffusion-weighted tractography. In this framework, each cortical region is represented as a neural mass governed by a mean-field equation describing the average synaptic gating variable S and its local dynamics. The structural connectome sets the anatomical coupling weights between regions. (**b**) Simulated activity generated whole-brain time series from which different data features were extracted using the same pipeline applied to empirical resting-state data. (**b**) Relationship between the global excitability parameter *w* and the global coupling *G* and representative dynamical features extracted from the simulated activity. Each point corresponds to a single simulation run performed with different parameter (*w, G*) values. Feature values were min-max normalized to the interval [0, 1] across all simulation priors.

The parameter *w* controls recurrent excitation uniformly across regions, whereas the global coupling parameter *G* scales interactions between regions. Representative simulated synaptic activity and empirical MEG signals are illustrated in Fig. 3b. To examine the contributions of these two mechanisms, we varied *w* and *G* jointly over the ranges [2, 4] and [1.5, 2.5], respectively. For each simulation, we computed the same six whole-brain ATM features used to characterize the empirical recordings: the mean and median of the upper and lower triangles, the Frobenius norm, and SVD-based entropy. Each feature was independently normalized across the simulation ensemble. The resulting parameter-space maps show how these observables depend on the combination of local recurrent excitation and global coupling (Fig. 3c-h). Mean and median transition probabilities generally increased with *w* in both triangles of the ATM, accompanied by an increase in the Frobenius norm. These changes indicate that stronger recurrent excitation enhances the propensity for avalanche-related activity to propagate across the network. Global coupling also modulated these features: at a given excitation level, differences in *G* shifted the resulting transition statistics. Thus, the magnitude of avalanche propagation reflected contributions from both local recurrent excitation and interactions mediated by structural connectivity. ATM entropy showed an opposing overall pattern, with lower values at higher recurrent excitation. Because this measure summarizes the distribution of normalized singular values, its reduction indicates that the transition matrix becomes more strongly dominated by a smaller number of components. Stronger propagation can therefore coexist with a more concentrated organization of avalanche-related interactions, consistent with the reinforcement of dominant configurations.

The direction of the simulated increases in the ATM mean, median, and norm, along with the positive ALS - CTRL effect sizes observed empirically, are consistent with hyperexcitability in ALS patents. The simulated decrease in entropy matched the direction of the (nonsignificant) empirical effect in the same direction. The dependence of the features on both *w* and *G* means that the empirical alterations cannot be attributed to recurrent excitation from these qualitative comparisons alone. We therefore used the simulated feature vectors to train a joint inference model for *p*(*w, G* | **y**), allowing us to estimate the recurrent excitation and the global coupling while also assessing the corresponding uncertainty.

Using simulations sampled from uniform priors over *w* ∈ [2, 4] and *G* ∈ [1.5, 2.5], we trained a neural density estimator to approximate the joint posterior *p*(*w, G* | **y**) (Fig. 4a). This analysis tested whether the two parameters could be distinguished despite their overlapping effects on large-scale activity propagation. Validation in 800 held-out synthetic simulations revealed different levels of recovery precision for the two parameters. The marginal posterior distributions, shown for six illustrative cases, successfully recovered the variation in ground-truth values (Fig. 4b,e). Across the entire test set, the posterior means of *w* closely followed the identity line, while the estimates of *G* exhibited greater dispersion (Fig. 4d,g). These results indicate that the selected ATM features supported the recovery of recurrent excitation, although the uncertainty is larger for *G* as compared to *w*. To assess the confidence of the inference, we additionally evaluated the posterior shrinkage and the posterior z-score metrics. According to established simulation-based inference diagnostics, the ideal inference regime is characterized by posterior shrinkage values approaching 1 and posterior z-scores close to 0, reflecting strong parameter identifiability and accurate posterior calibration [44, 45]. In our analysis, shrinkage was generally higher for *w* than for *G*, indicating a stronger reduction in uncertainty relative to the respective priors (Fig. 4c,f) and demonstrating that the observed features strongly constrained the latent parameter space. Absolute posterior z-scores were low for most test cases, although larger standardized errors occurred for both parameters. Additional parameter-dependent recovery analyses revealed that estimation accuracy and credible-interval coverage varied across the investigated prior ranges, with larger errors and reduced coverage near some parameter’s boundaries (Supplementary Figs. SM3 and SM4). These results indicate that aggregate recovery statistics may conceal local estimation biases and nonuniform posterior calibration, which should be considered when interpreting individual parameter estimates. Together, these findings support the feasibility of jointly estimating recurrent excitation and global coupling from whole-brain ATM features, with stronger relative posterior concentration and more accurate point recovery for *w*. The synthetic validation provides a basis for applying the joint model to empirical recordings and examining whether the ALS vs CTRL difference in recurrent excitation persists when global coupling is estimated rather than fixed.

**Figure 4:**
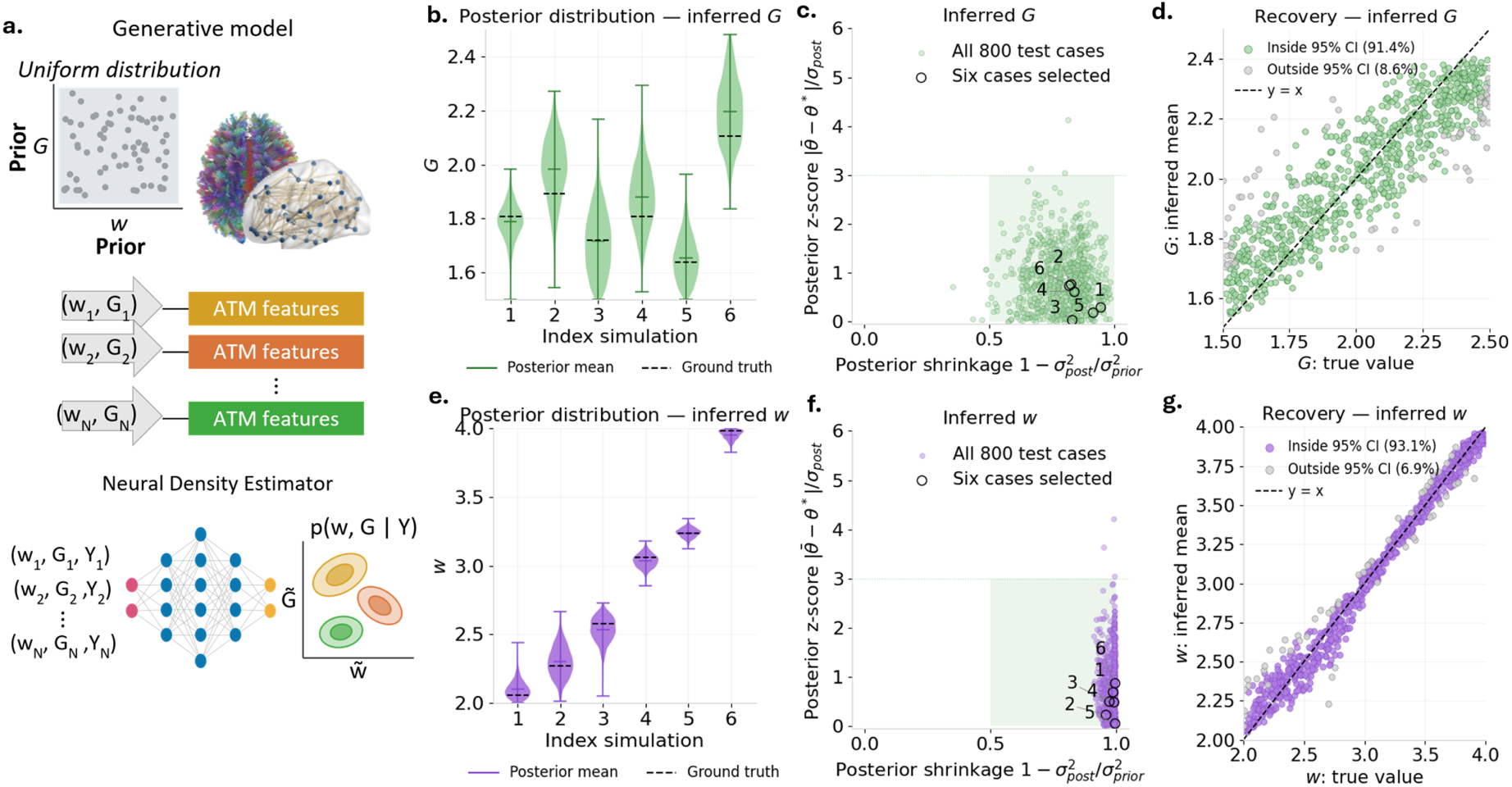
Synthetic validation of joint inference of recurrent excitability and global coupling. **(a)** Inference workflow. Parameter pairs (*w, G*) were sampled from uniform priors, and whole-brain ATM features were extracted from the resulting simulations. A neural density estimator was trained to approximate the joint posterior *p*(*w, G |* **y**). **(b,e)** Marginal posterior distributions of *G* (green) **(b)** and *w* (purple) **(e)** for six illustrative test simulations. Matching indices identify the same simulation across panels. Colored horizontal lines indicate posterior means, and dashed horizontal lines indicate the true parameter values used to generate the synthetic data, demonstrating accurate posterior recovery and low estimation bias. **(c,f)** Posterior shrinkage, 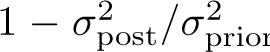, versus absolute posterior z-score, |*θ̄*−*θ^∗^*| */σ*_post_, for all 800 held-out test simulations. Black outlined circles identify the six illustrative cases. The shaded area highlights high shrinkage and relatively low standardized estimation error. **(d,g)** Posterior means plotted against ground-truth values for *G* and *w*, respectively. The dashed line denotes perfect recovery. Colored points indicate cases in which the ground-truth value falls within the marginal 95% credible interval; gray points indicate cases outside that interval. Empirical coverage was 91.4% for *G* and 93.1% for *w*. This indicates that the inversion pipeline provides accurate, identifiable, and well-constrained estimates of the global excitability parameter from whole-brain dynamical features.

### 2.3 Bayesian estimation of cortical excitability reveals hyperexcitability in ALS

We next applied the validated inference framework to empirical whole-brain ATM features, jointly estimating the global recurrent-excitation parameter *w* and the global coupling *G* for each participant. The parameter *w* modulates the effective recurrent excitation of cortical populations and therefore controls the operating regime of large-scale brain dynamics. This approach allowed us to assess whether the ALS-related increase in recurrent excitation persisted when interregional coupling was estimated rather than fixed. Figure 5a,b shows the joint posterior distributions for illustrative (i.e., selected randomly) ALS and control participants. The elongated contours indicate posterior dependencies between *w* and *G*, highlighting the importance of retaining uncertainty in both parameters during model inversion. These individual examples illustrate the joint estimates; group comparisons were based on subject-level posterior means. The inferred global coupling did not differ significantly between groups (*p* = 0.558, *d* = 0.14; Fig. 5c). In contrast, inferred recurrent excitation was significantly higher in ALS patients than in controls (two-sided Welch’s *t*-test, *p* = 0.00432, Cohen’s *d* = 0.68; Fig. 5e). Although the distributions overlapped, ALS patients showed a group-level shift toward higher *w* values. Thus, the increase in model-derived recurrent excitation remained detectable when global coupling was allowed to vary across participants. Within the tested model and prior ranges, this finding supports an interpretation of the empirical ALS dynamics in terms of enhanced recurrent excitation, rather than an effect contingent on fixing *G* to a single value. The absence of a statistically significant group difference in *G* suggests that the observed alterations are more strongly associated with recurrent excitation within the present model. All in all, these results indicate that the empirical dynamical alterations observed in ALS are consistent with a shift toward a hyperexcitable cortical state.

**Figure 5:**
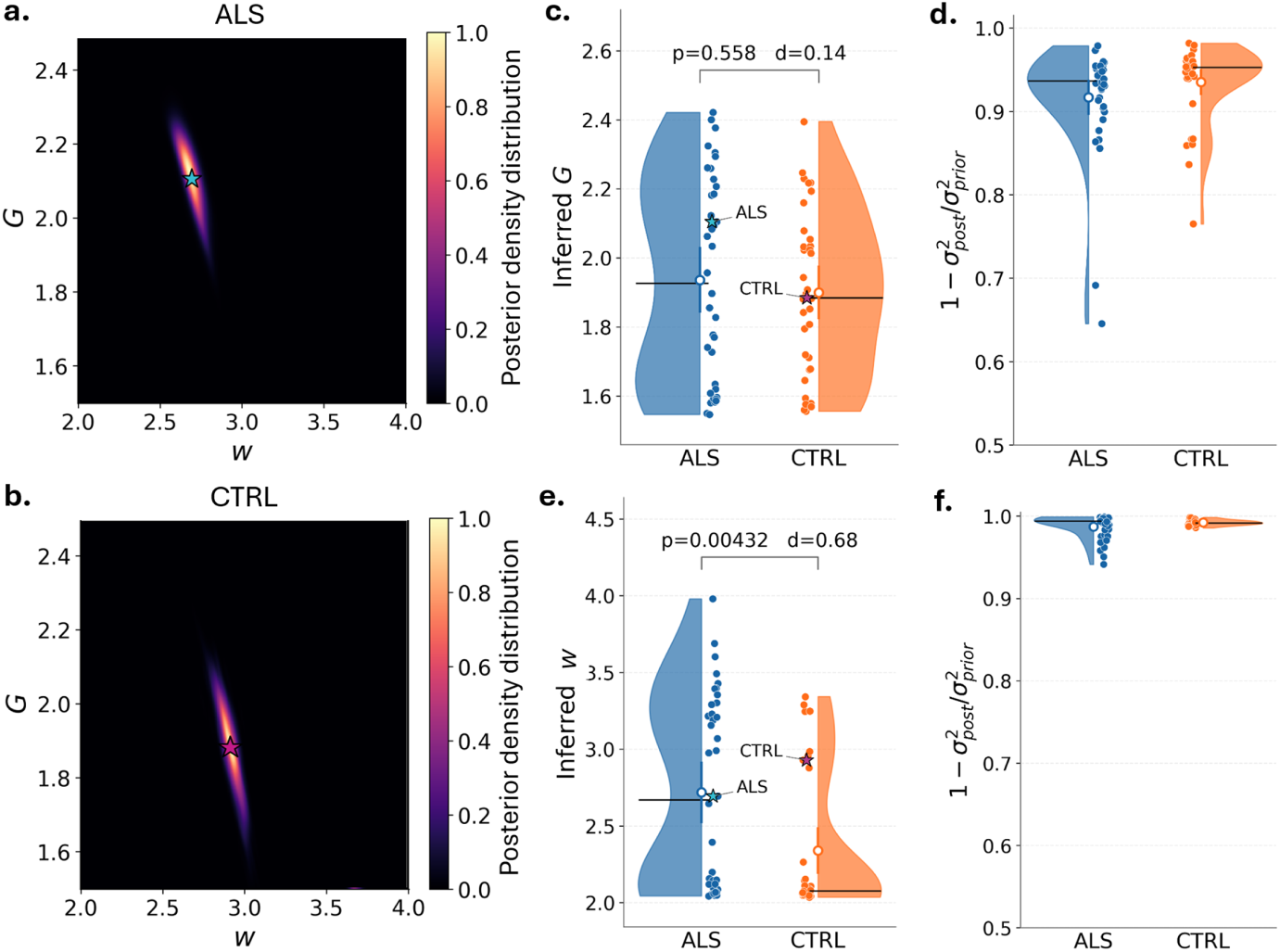
Joint Bayesian inference of global recurrent excitation and coupling in ALS patients and healthy controls. **(a,b)** Joint posterior density maps for illustrative ALS and control participants, respectively, showing the inferred global recurrent-excitability parameter *w* (horizontal axis) and global coupling *G* (vertical axis). Color indicates relative posterior density. Star markers identify the mean value of w and G. **(c,d)** Distributions of subject-level posterior means for *G* and its subject-level posterior shrinkage, 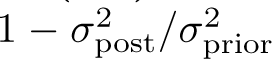, respectively, in ALS patients (blue) and controls (orange). Each point represents one participant. Global coupling did not differ significantly between groups (two-sided Welch’s *t*-test, *p* = 0.558, Cohen’s *d* = 0.14). The shrinkage Values approaching one indicate a substantial reduction in parameter uncertainty relative to the prior. **(e,f)** Distributions of subject-level posterior means for *w* and its subject-level posterior shrinkage, 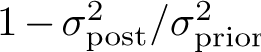, respectively, as before. Inferred recurrent excitability was higher in ALS patients (*p* = 0.00432, *d* = 0.68). ALS patients show significantly elevated *w_G_* values relative to controls, supporting the hypothesis of globally increased cortical excitability underlying altered large-scale brain dynamics in ALS.

To assess the reliability and informativeness of the inversion procedure, we quantified posterior shrinkage, which measures how much the empirical data constrain the posterior distribution relative to the prior distribution. Posterior shrinkage indicated substantial reductions in uncertainty relative to the priors for both parameters (Fig. 5d,f). Shrinkage for *w* was consistently close to one, whereas *G* exhibited greater variability across participants, consistent with the differential precision observed in synthetic validation. These results show that the fitted posterior distributions were more concentrated than the priors, particularly for recurrent excitation.

Together, these results indicate that ALS-related changes in large-scale avalanche dynamics are compatible with increased model-derived recurrent excitation, even when uncertainty in global coupling is explicitly incorporated. This finding supports recurrent excitation as a candidate mechanism contributing to the altered propagation dynamics observed in ALS, while remaining distinct from an independent physiological measurement of cortical excitability.

### 2.4 Effects of regional recurrent excitation on whole-brain dynamics

Building on the spatially uniform model, we next investigated regional differences in recurrent excitation. We distinguished excitation in the 34 motor-frontal regions (*w*_F_) from that in the remaining 46 regions (*w*_NF_). In this configuration, both regional excitation parameters and global coupling *G* were inferred jointly from the whole-brain ATM features. To illustrate the effects of regional excitation, Figure 6 shows an exploratory parameter-space analysis at a fixed illustrative coupling value of *G* = 1.89. This value corresponds to the mean of the subject-specific posterior estimates of *G* obtained for healthy controls in the preceding spatially uniform model inversion. For each pair (*w*_F_*, w*_NF_), we generated regional time series and computed whole-brain activity summaries (that is, the average synaptic activity and its variance) and the six ATM features extracted from the empirical recordings. The fixed coupling value was used only for this exploratory analysis; global coupling remained a free parameter in the subsequent subject-specific inference. Fig. 6a shows higher mean synaptic activity toward parameter combinations with high excitation in both partitions, along with decreased variance. Stronger recurrent excitation was therefore associated with higher sustained activity and smaller fluctuations. Representative frontal time series from the highlighted parameter combinations illustrate these contrasting regimes (Fig. 6b). Within the range *w*_F_*, w*_NF_ ∈ [2, 4], the six whole-brain ATM features exhibited distinct dependencies on the two parameters (Fig. 6c-h). Each point represents one simulation, with color indicating the corresponding feature value normalized across the displayed simulation ensemble. Mean and median transition probabilities generally increased with recurrent excitation (Fig. 6c,d,e,f). Both parameters nevertheless contributed to the resulting feature landscapes. The Frobenius norm increased toward combinations with high excitation in both partitions (Fig. 6g), consistent with an increased overall magnitude of avalanche-related transition probabilities. ATM entropy showed the opposite overall pattern, with the lowest values occurring when excitation was high in both partitions (Fig. 6h). Together with the reduction in synaptic-activity variance, this pattern is compatible with a regime in which stronger activity propagation coexists with more constrained dynamics. Increased excitation may therefore reinforce dominant configurations of coordinated activity rather than expand the diversity of configurations explored by the network. The differing sensitivities of the whole-brain features motivated their combined use for joint inference of (*G, w*_F_*, w*_NF_). This formulation allowed us to investigate regional differences in recurrent excitation while accounting for uncertainty in global coupling and retaining posterior dependencies among all three parameters.

**Figure 6:**
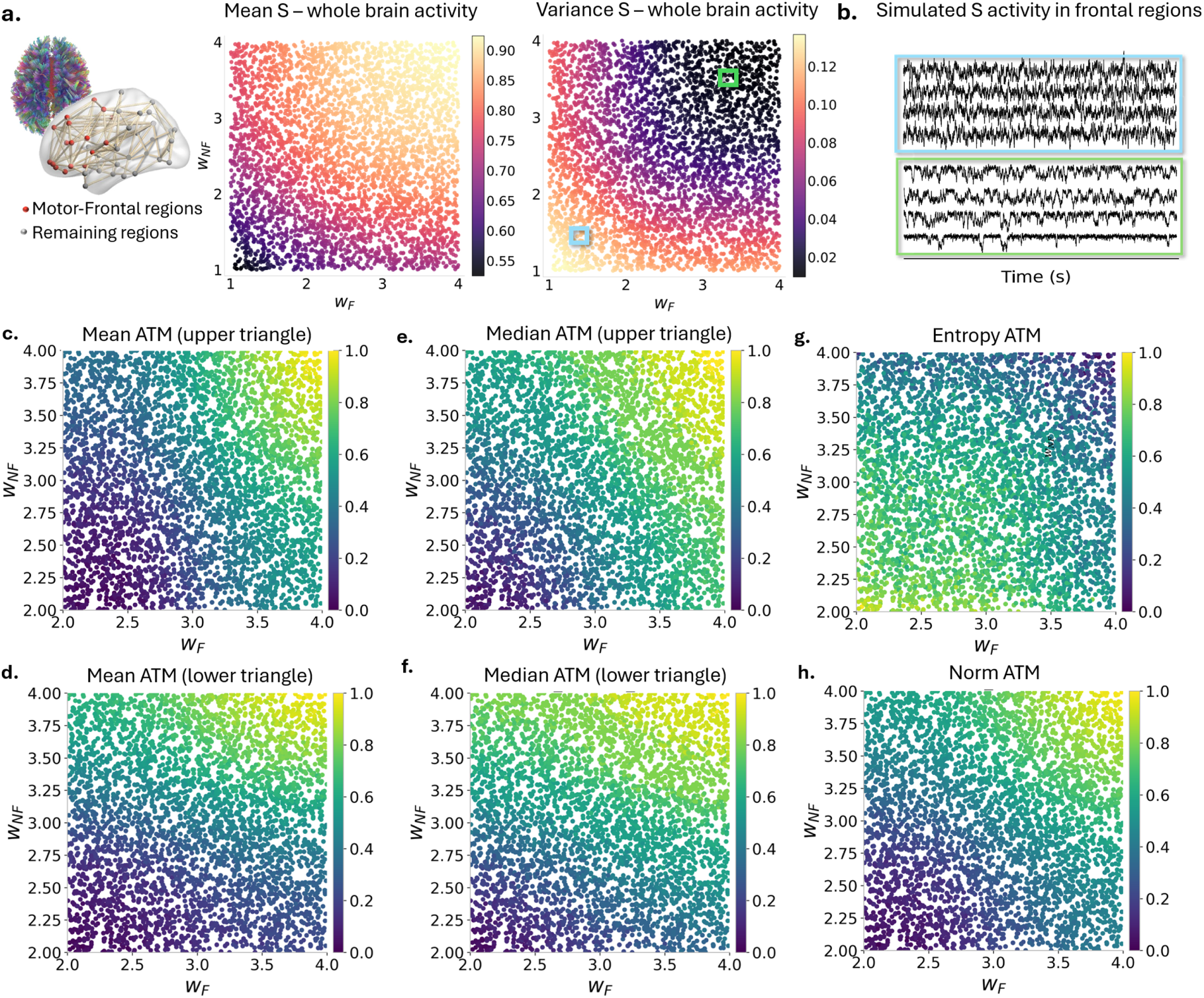
Effects of regional recurrent excitation on whole-brain activity and avalanche transition features. Two recurrent excitability parameters were varied independently: *w*_F_, assigned to the 34 motor-frontal regions, and *w*_NF_, assigned to the remaining 46 regions. This figure illustrates how variations in regional recurrent excitation affect whole-brain dynamics and avalanche transition features. For this illustrative exploration, global coupling was fixed at *G* = 1.89, corresponding to the mean of the control-group posterior distribution obtained from the initial joint inversion of global recurrent excitation and coupling. Each point represents a simulation with a fixed pair of *w*_F_ and *w*_NF_. This illustrative exploration is distinct from the subsequent subject-specific inversion, in which *w*_F_, *w*_NF_, and *G* were jointly inferred for each participant. Each point represents a simulation with a fixed parameter pair. **(a)** Mean synaptic activity ⟨*S*⟩ and its variance across the parameter space *w*_F_*, w*_NF_ ∈ [1, 4]. The horizontal and vertical axes indicate *w*_F_ and *w*_NF_, respectively; the color represents the activity statistic. Colored squares identify the parameter combinations used for the example time series. **(b)** Representative simulated synaptic activity in frontal regions for the combinations highlighted in (a), with matching border colors. **(c-h)** Whole-brain ATM features over *w*_F_*, w*_NF_ ∈ [2, 4]: upper- and lower-triangle means (c,d), upper- and lower-triangle medians (e,f), Frobenius norm (g), and SVD-based entropy (h). Color indicates each feature’s independently min-max-normalized value. The norm generally increases and SVD-based entropy decreases toward parameter combinations with high excitability in both partitions.

### 2.5 Joint regional inference identifies elevated motor-frontal recurrent excitability associated with functional impairment

We next applied the regional model to empirical whole-brain ATM features, jointly estimating motor-frontal recurrent excitation *w*_F_, other regions recurrent excitation *w*_NF_, and global coupling *G* for each participant. This analysis investigated whether allowing recurrent excitation to vary between anatomical partitions revealed alterations and clinical associations not captured by the spatially uniform model.

To infer the three parameters from empirical observations, we employed the same simulation-based inference framework used for the spatially uniform model. Specifically, a neural density estimator was trained on simulated whole-brain dynamics generated from known combinations of (*G, w*_F_*, w*_NF_), learning an approximation of the joint posterior distribution conditioned on the six whole-brain ATM features. This approach enabled simultaneous estimation of the two regional recurrent-excitation parameters and global coupling while retaining potential dependencies among them, similarly to previous applications of Bayesian whole-brain model inversion [40].

Before applying the trained inference framework to empirical observations, we assessed its ability to recover known model parameters using held-out synthetic simulations excluded from training. Fig. 7 presents the results for global coupling *G* (a-c), motor-frontal recurrent excitation *w*_F_ (d-f), and non-frontal recurrent excitation *w*_NF_ (g-i). respectively, for six illustrative simulations. The posterior means were compared with the corresponding ground-truth parameter values to assess the accuracy and uncertainty of the inferred estimates. To quantify parameter recovery across the entire range of the test ensemble, we examined posterior shrinkage as a function of the absolute standardized estimation error (Fig. 7b,e,h). Posterior shrinkage measures the reduction in uncertainty relative to the prior, whereas the standardized estimation error quantifies the discrepancy between the posterior mean and the ground-truth value relative to posterior uncertainty. These complementary diagnostics allow us to assess whether concentrated posterior distributions are also associated with accurate parameter estimates. Fig. 7c,f, i further compares posterior mean estimates with their corresponding ground-truth values. Overall, the inferred parameters followed the identity relationship, supporting the ability of the framework to recover regional recurrent-excitation parameters from whole-brain dynamical features. Overall, inferred parameter values followed the identity relationship, although recovery accuracy varied across the parameter space. The recurrent-excitation parameters showed relatively concentrated recovery patterns, whereas global coupling exhibited greater dispersion around the identity line.

**Figure 7:**
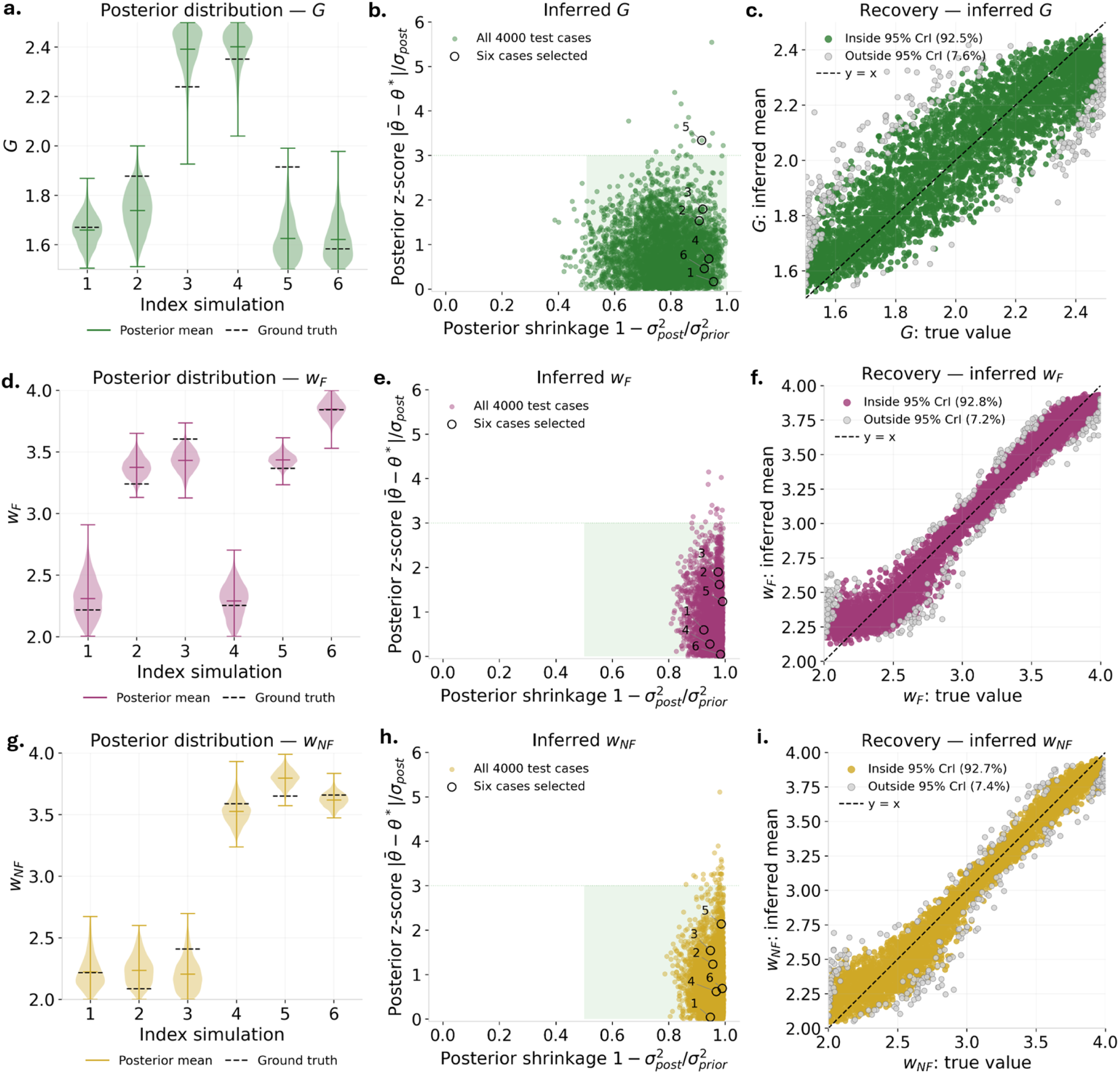
Synthetic validation of joint inference of motor-frontal recurrent excitation, non-frontal recurrent excitation, and global coupling. The three parameters were inferred jointly from whole-brain ATM features. **(a,d,g)** Marginal posterior distributions of *G* (green), *w*_F_ (purple), and *w*_NF_ (yellow), respectively, for six illustrative held-out test simulations. Matching indices identify the same simulation across panels. Violin widths represent posterior density, colored horizontal lines indicate posterior means, and black dashed lines mark the corresponding ground-truth values. **(b,e,h)** Posterior shrinkage, defined as 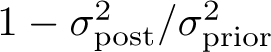, versus absolute posterior z-score, |*θ̄* − *θ^∗^*|*/σ*_post_, for the held-out simulations. Here, *θ̄* denotes the posterior mean and *θ^∗^* the ground-truth parameter. Black outlined circles identify the six cases shown in panels (a,d,g). The shaded area highlights high shrinkage and relatively low standardized estimation error. Most estimates fall within this region, demonstrating that the inversion is both accurate and well-calibrated, without major overconfidence or bias. **(c,f,i)** Posterior mean estimates are plotted against the corresponding ground-truth values for *G*, *w*_F_, and *w*_NF_, respectively. The black dashed line indicates perfect recovery (*y* = *x*). Colored points represent simulations for which the ground-truth parameter falls within the marginal 95% interval. The results demonstrate informative recovery of all three parameters, with greater variability for .global coupling and slightly more uncertain parameter recovery for low parameter values.

Having assessed parameter recovery on synthetic data, we subsequently applied the trained joint inference framework to empirical whole-brain ATM features to estimate subject-specific posterior distributions of (*G, w*_F_*, w*_NF_).

Figure 8a,b shows marginal and pairwise posterior distributions for illustrative ALS and control participants. Group comparisons were performed using the posterior mean of each participant. ALS patients exhibited higher motor-frontal excitation than controls (two-sided Welch’s *t*-test, *p* = 5.69×10*^−^*^5^, Cohen’s *d* = 1.00; Fig. 8e). Non-frontal excitation was also higher in ALS, with a smaller effect size (*p* = 0.00727, *d* = 0.64; Fig. 8g). Global coupling did not differ significantly between groups (*p* = 0.872, *d* = 0.04; Fig. 8c). These results indicate that increased model-derived recurrent excitation is not restricted to motor-frontal regions. Nevertheless, the larger standardized group effect observed for *w_F_* is consistent with a more pronounced alteration in the motor-frontal partition. Posterior shrinkage indicated substantial concentration relative to the priors for all three parameters, particularly for the regional excitation parameters (Fig. 8d,f,h). The subject-level parameter plane illustrates heterogeneity in the relative excitation of the two partitions (Fig. 8i). Posterior means satisfied *w̄*_F_ *> w̄*_NF_ in 29 of 38 ALS patients (76.3%) and 24 of 35 controls (68.6%). These proportions describe the ordering of the estimates rather than establishing a disease-specific regional imbalance.

**Figure 8:**
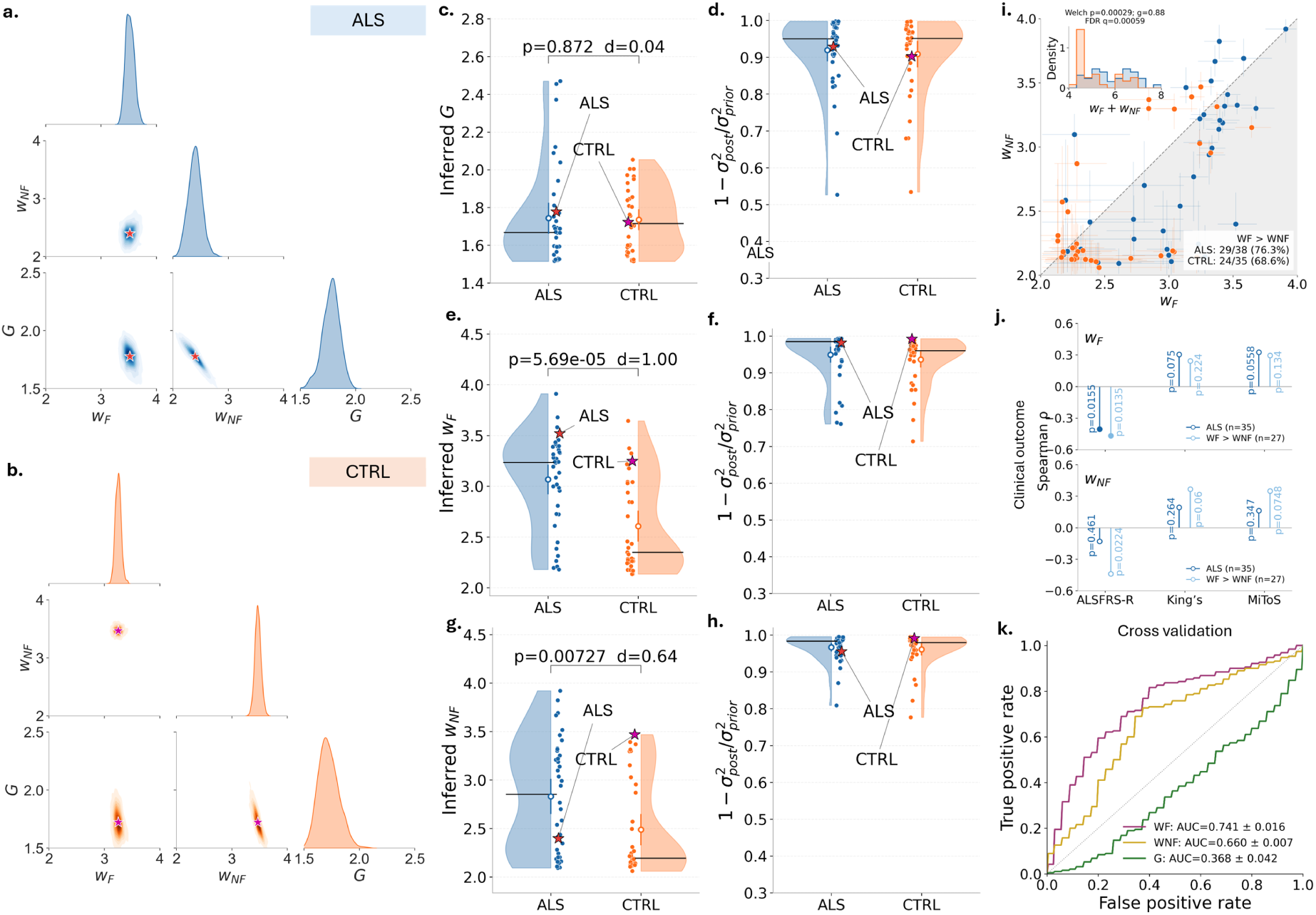
Subject-specific Bayesian inference of motor-frontal and non-frontal recurrent excitability in ALS patients and healthy controls. (a,b) Posterior distributions for two illustrative participants, one with ALS (blue) and one control (orange). Diagonal panels show marginal posterior densities of *w*_F_, *w*_NF_, and *G*; off-diagonal panels show their pairwise posterior densities. Stars mark the coordinates defined by the corresponding posterior means. **(c,e,g)** Distributions of subject-level posterior means for *G*, *w*_F_, and *w*_NF_, respectively, in ALS patients (blue, *n* = 38) and controls (orange, *n* = 35). Each point represents one participant. Global coupling did not differ significantly between groups (two-sided Welch’s *t*-test, *p* = 0.872, Cohen’s *d* = 0.04). ALS patients exhibited higher estimates of both regional excitability parameters, with a larger effect size for *w*_F_ (*p* = 5.69 10*^−^*^5^, *d* = 1.00) than for *w*_NF_ (*p* = 0.00727, *d* = 0.64). Stars highlight the same illustrative participants shown in panels (a,b). **(d,f,h)** Subject-level posterior shrinkage, 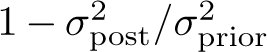, for *G*, *w*_F_, and *w*_NF_, respectively. Values approaching one indicate substantial posterior concentration relative to the corresponding prior. Stars identify the illustrative participants. **(i)** Subject-level posterior means in the (*w*_F_*, w*_NF_) plane, with bars indicating marginal posterior uncertainty. The dashed identity line separates participants with *w*_F_ *> w*_NF_ (below) from those with *w*_NF_ *> w*_F_ (above). The shaded region highlights *w*_F_ *> w*_NF_, observed in 29 of 38 ALS patients (76.3%) and 24 of 35 control s (68.6%). The inset shows the distributions of the sum of subject-level posterior means, *w̄*_F_ + *w̄*_NF_, which summarizes the combined level of the two regional excitability parameters without weighting by partition size. The group comparison for this sum is reported above the inset (Welch’s *p* = 0.00029, Hedges’ *g* = 0.88, FDR-adjusted *q* = 0.00059). **(j)** Spearman rank-order correlations between the posterior means of *w*_F_ (top) and *w*_NF_ (bottom) and clinical outcomes: ALSFRS-R, King’s stage, and MiToS stage. Correlations are shown for the ALS sample included in the clinical analysis (*n* = 35, dark blue) and for the subgroup with *w*_F_ *> w*_NF_ (*n* = 27, light blue). Filled circles indicate correlations that remain significant after false discovery rate correction (FDR-adjusted *p <* 0.05); open circles indicate correlations that do not survive this correction. **(k)** Receiver operating characteristic curves for ALS–control classification using each inferred parameter separately, evaluated by nested five-fold cross-validation repeated five times. Mean AUC ± standard deviation across repetitions was 0.741 ±0.016 for *w*_F_, 0.660 ±0.007 for *w*_NF_, and 0.368 ±0.042 for *G*. The dashed diagonal indicates chance-level discrimination.

The unweighted sum of the two posterior means, *w̄_F_*+ *w̄_NF_* , was also higher in ALS patients than in controls (Welch’s *p* = 0.00029, FDR-adjusted *q* = 0.00059, Hedges’ *g* = 0.88; Fig. 8i, inset), consistent with an overall increase in model-derived recurrent excitation.

We then assessed clinical associations in the ALS sample available for this analysis (*n* = 35; Fig. 8j). Higher motor-frontal excitation was associated with lower ALSFRS-R scores, indicating greater functional impairment. This association survived the specified FDR correction (*ρ* = −0.406*, p* = 0.0155*, q* = 0.0464). Associations with King’s and MiToS stages did not survive correction, and no corrected significant associations were observed for *w*_NF_ and for *G*. Unlike the spatially uniform excitation parameter, motor-frontal excitation therefore showed a detectable association with functional impairment. These findings suggest that the regional distribution of recurrent excitation may contain clinically relevant information not fully captured by a spatially uniform parameterization. We evaluated ALS-CTRL discrimination using each parameter separately (Fig. 8k). Nested five-fold cross-validation repeated five times yielded mean areas under the receiver operating characteristic curve of 0.741 ± 0.016 for *w*_F_, 0.660 ± 0.007 for *w*_NF_, and 0.368 ± 0.042 for *G*, where the reported variation is the standard deviation across repetitions. Motor-frontal excitation achieved the highest mean AUC among the individual parameters. As an exploratory extension, we evaluated the sum and difference of the inferred regional weights (Supplementary Fig. SM5). The sum (*w_F_* + *w_NF_*) achieved an AUC of (0.735 ± 0.010), numerically close to that obtained with *w_F_* alone (0.741 ± 0.016), whereas the difference (*w_F_* − *w_NF_*) showed lower discrimination (0.563 ± 0.025). In label-permutation tests, the sum performed above the permutation null (Holm-adjusted (*p* = 0.005), whereas the difference did not (adjusted (*p* = 0.390). To assess whether model inversion preserved or enhanced disease-related information contained in the empirical observations, we compared the discrimination obtained using the six original ATM features with that obtained using the inferred parameters. Under the same nested cross-validation procedure, the six empirical ATM features achieved an AUC of (0.748 ± 0.029), compared with (0.741 ± 0.016) for motor-frontal recurrent excitation alone and (0.700 ± 0.025) for the three jointly inferred parameters, as depicted in Fig.SM5 of Supplementary Material. Thus, the single motor-frontal parameter retained a discriminatory performance numerically close to that obtained from the full empirical feature set.

### Subject-level simulations and empirical feature comparison

To assess whether the inferred parameters reproduced the empirical data features, ten stochastic simulations were generated for each of the 73 non-outlier subjects (38 ALS and 35 controls), starting from the posterior means of *G*, *w_F_* and *w_NF_* . The six ATM features were computed using the same feature extraction procedure employed for the simulation ensemble. The newly simulated features were transformed using the original simulation min-max scaler. The empirical features were transformed using the original empirical scaler, shared across ALS and controls. Thus, each subject contributed one empirical and one simulated value per feature in the normalized representation used for inference. For each feature and diagnostic group, empirical and simulated values were displayed as adjacent box-and-whisker plots. Boxes represented the interquartile range, with the median indicated inside each box; whiskers extended to the most extreme observations within 1.5 interquartile ranges of the box boundaries. For each subject, ten stochastic simulations were generated using the posterior mean estimates of *G*, *w_F_* and *w_NF_* , with independent noise realizations. For each feature, the median across the ten simulations was computed to obtain a single representative simulated value per subject. Differences between the empirical and simulated marginal distributions were assessed using a two-sample Cramér-von Mises test, without using subject pairing in the test statistic. The resulting *p*-values were adjusted using the Benjamini-Hochberg procedure across the six features within each diagnostic group. Significance annotations on the boxplots refer to these adjusted values. Subject-level correspondence was assessed separately using Spearman correlations between empirical and simulated values for each feature within each group. For an additional visualization in empirical units, normalized simulated values were transformed using the inverse empirical scaler. This remapping preserved their normalized discrepancies and did not represent the raw feature values produced by the simulator. Accordingly, agreement in the normalized or remapped representation was distinguished from agreement between raw empirical and simulated features. To further characterize the dynamical organization of the simulated activity, we compared neuronal avalanche-size and duration distributions between empirical recordings and subject-specific simulations (Supplementary Fig.SM6). Simulations were generated using the inferred posterior mean estimates of *w_F_* , *w_NF_* , and *G* for each participant. Both empirical and simulated signals exhibited broad avalanche-size and duration distributions with qualitatively similar decreasing profiles. Nevertheless, quantitative differences emerged in their scaling exponents. These results indicate that the model generates avalanche-like activity with qualitative similarities to empirical recordings, although it does not fully reproduce their statistical distributions. Importantly, avalanche-size and duration distributions were not directly included among the features used for model inversion, making these comparisons complementary assessments of the simulated dynamics.

## 3 Discussion

In this study, we combined source-reconstructed MEG recordings, whole-brain dynamical modeling, and simulation-based inference to investigate whether alterations in large-scale brain dynamics in amyotrophic lateral sclerosis (ALS) could be explained by changes in recurrent excitation. Building on previous analyses of this cohort [20] and established whole-brain modeling approaches [27,28], we first characterized the empirical dynamics using directed avalanche transition matrices (ATMs). We then examined how recurrent excitation and interregional coupling shaped the corresponding observables in a reduced Wong–Wang model. Finally, we inferred subject-specific model parameters using two complementary configurations: a spatially uniform model and a regional model distinguishing motor-frontal from the remaining brain regions. The motor-frontal partition allowed us to examine whether whole-brain avalanche dynamics were compatible with preferential recurrent excitation in motor and frontal systems, motivated by their involvement in ALS. This partition is a hypothesis-driven simplification of the disease’s heterogeneous anatomical distribution. In particular, it does not explicitly separate frontotemporal systems associated with cognitive and behavioral manifestations, which may occur independently of memory impairment. Alternative partitions distinguishing extended motor, frontotemporal, and remaining regions could provide a more differentiated account, but would require additional validation of parameter identifiability and more refined clinical stratification, especially in terms of assessments of extra-motor symptoms. The results indicate that ALS-related changes in avalanche propagation are consistent with increased model-derived recurrent excitation, while the regional parameterization reveals an association between motor-frontal excitation and functional impairment.

The empirical analysis revealed higher avalanche transition probabilities in ALS patients than in healthy controls, particularly in the mean and median of the upper ATM triangle, together with an increased Frobenius norm (Fig.2). These findings indicate an increased propensity for sequential regional recruitment during neuronal avalanches. Importantly, the ATM describes the temporal ordering of suprathreshold activations within avalanche events, rather than direct anatomical or causal interactions. The different effect magnitudes observed in the upper and lower triangular summaries reflect properties of the directed transition structure under the chosen regional ordering and should not be interpreted as evidence for alterations in distinct anatomical pathways. In contrast to the measures of transition magnitude, SVD-based ATM entropy showed a negative but nonsignificant group difference. Although stronger propagation could coexist with a greater concentration of activity in dominant dynamical configurations, the present results do not establish a reduction in the diversity or complexity of brain dynamics in ALS. ATM-derived spectral entropy should be distinguished from sequence-based measures of neural complexity, such as Lempel–Ziv complexity. While the latter quantifies the diversity and compressibility of temporal or spatiotemporal activity sequences, ATM spectral entropy characterizes the distribution of singular-value weight in a matrix summarizing interregional avalanche propagation. Accordingly, these measures capture different properties of neural dynamics and need not exhibit the same relationship with cortical excitability. In fact, some models suggest that excitability would relate to higher complexity [46], while others point in the opposite direction. In our simulations, increased recurrent excitation was associated with stronger avalanche propagation and lower ATM spectral entropy, suggesting a greater concentration of transition structure into dominant matrix components. However, the empirical reduction in ATM entropy observed in ALS patients did not reach statistical significance. Therefore, our findings do not establish a reduction in temporal neural complexity in ALS. Overall, the most consistent empirical finding was an increase in avalanche-related transition strength. The modeling results provide a mechanistic bridge between the empirical alterations observed in ALS and recurrent excitation, which is not directly observable from the MEG features, and is understood here as a hidden variable. Hence, the model’s epistemic scope accounts to providing a possible mechanistic interpretation of these observations. Increasing local recurrent excitation produced higher ATM mean and median values and a larger Frobenius norm, reproducing the direction of the principal empirical differences between ALS patients and controls (Fig. 3). The simulated decrease in ATM entropy was also directionally consistent with the empirical effect, although the latter was not statistically significant. These relationships suggest that stronger local recurrent feedback can promote sequential activity propagation across structurally connected regions. However, global coupling also influenced the simulated features, indicating that recurrent excitation and interregional interactions jointly determine the resulting whole-brain dynamics. This motivated the joint inference of excitation and coupling parameters, rather than attributing the observed alterations to recurrent excitation based on parameter-space explorations alone.

To infer latent model parameters from empirical data, we used simulation-based inference for Bayesian model inversion. This approach is well suited to whole-brain neural mass models, where the relationship between parameters and observed dynamics is nonlinear, stochastic, and analytically intractable. Simulations across the prior parameter ranges were used to train neural density estimators of subject-specific posterior distributions. Validation on held-out synthetic simulations supported parameter recovery, although precision varied across parameters. Posterior shrinkage further indicated substantial reductions in uncertainty relative to the priors, particularly for excitation parameters. The two inference configurations addressed complementary questions. In the first, recurrent excitation was assumed spatially uniform, and (*w, G*) were inferred jointly. In this case, ALS patients exhibited higher inferred recurrent excitation than controls, whereas global coupling did not differ significantly between groups (Fig. 5). The persistence of the excitation effect when coupling was estimated simultaneously supports the interpretation that the observed dynamical alterations are compatible with increased recurrent excitation within the investigated model and parameter ranges. Nevertheless, the absence of a significant group difference in coupling does not demonstrate equivalence between groups, and posterior dependencies indicate that compensatory relationships between parameters remain possible. Accordingly, the inferred excitation parameter should be understood as an effective model-derived quantity describing local recurrent feedback under the assumed generative model, rather than a direct measurement of physiological cortical excitability. As such, the inferred parameters will require empirical validation in future studies. This analysis tested whether ALS-related dynamics were compatible with an overall increase in excitation while accounting for global coupling. Although inferred *w* distinguished ALS patients from controls, it did not show statistically significant associations with the clinical measures examined.

The regional configuration, jointly inferring (*G, w*_F_*, w*_NF_), examined whether the spatial distribution of excitation carried clinical information not captured by a single uniform parameter. Both motor-frontal (*w_F_*) and remaining-region *w_NF_* recurrent-excitation parameters were higher in ALS patients, with a numerically larger standardized group effect for (*w_F_*) (Fig. 8). The combined elevation of the two parameters was supported by the significant group difference in their sum. However, the regional contrast (*w_F_* − *w_NF_*) did not differ significantly between groups. Thus, although the observed effect was more pronounced for motor-frontal excitation, the results do not establish a statistically significant disease-specific imbalance between the two anatomical partitions. The findings support a distributed increase in model-derived recurrent excitation, with a potentially greater motor-frontal contribution that requires further investigation. This distinction is particularly important because the motor-frontal partition includes both motor-related and frontal association regions and does not represent an exclusively motor network. Moreover, both regional parameters were inferred from whole-brain ATM summaries, making their anatomical interpretation dependent on the predefined partition and the assumptions of the model. The regional parameterization revealed an association with clinical impairment. Higher motor-frontal recurrent excitation was associated with lower ALSFRS-R scores, and this association survived correction for multiple comparisons (Fig. 8j). In contrast, the spatially uniform excitation parameter did not show a corrected significant clinical association (Supplementary Fig. SM2), nor did the individual empirical ATM features (Supplementary Fig. SM1). This suggests that, although the overall elevation of recurrent excitation may characterize ALS-related alterations in large-scale dynamics, the motor-frontal component may provide additional information about interindividual differences in functional impairment. Such an interpretation remains exploratory and does not establish that motor-frontal excitation is more strongly associated with clinical impairment than the other inferred parameters. For these reasons, the model inversion may integrate information distributed across several dynamical observables into a compact parameter associated with functional impairment.

The classification analyses further clarify the contribution of the inferred parameters. Motor-frontal excitation achieved a mean cross-validated AUC of (0.741 ± 0.016), numerically close to the (0.748 ± 0.029) obtained using the six original empirical ATM features, whereas the three jointly inferred regional parameters achieved an AUC of (0.700±0.025) (Fig. 8k; Supplementary Fig. SM3). These findings indicate that a single inferred parameter retained much of the disease-discriminative information represented by the original feature set, without demonstrating superior classification performance. Because the inferred parameters are estimated from the empirical observables, they cannot be interpreted as providing independent information absent from those inputs. Their principal contribution lies instead in translating descriptive properties of large-scale activity into parameters with a defined role in a generative dynamical model. Unlike the original ATM summaries, these parameters can be systematically varied to investigate the consequences of altered recurrent excitation and coupling, including dynamical regimes not directly sampled in the empirical recordings. Such simulations provide a basis for formulating mechanistic hypotheses and potentially testable predictions, rather than establishing diagnostic or clinical predictive utility, which will require perspective validation.

We also evaluated the generative model against empirical observations by producing subject-specific simulations using the inferred posterior mean parameters. For each participant, ten stochastic realizations were generated and the resulting ATM features were compared with those extracted from MEG recordings (Fig. 9). These analyses assessed whether the inferred parameter configurations could generate dynamical summaries compatible with the observations and characterized variability across noise realizations. The distribution of the empirical and simulated features were overlapping. However, one should keep in mind that the inference had been performed utilizing normalized data and, as such, the simulations also underent the respective anti-transformation. Hence, the similarity of the distributions should not be interpreted in terms of raw , but rather relative, magnitudes. These analyses therefore constitute a model-fit assessment rather than complete posterior predictive validation.

**Figure 9:**
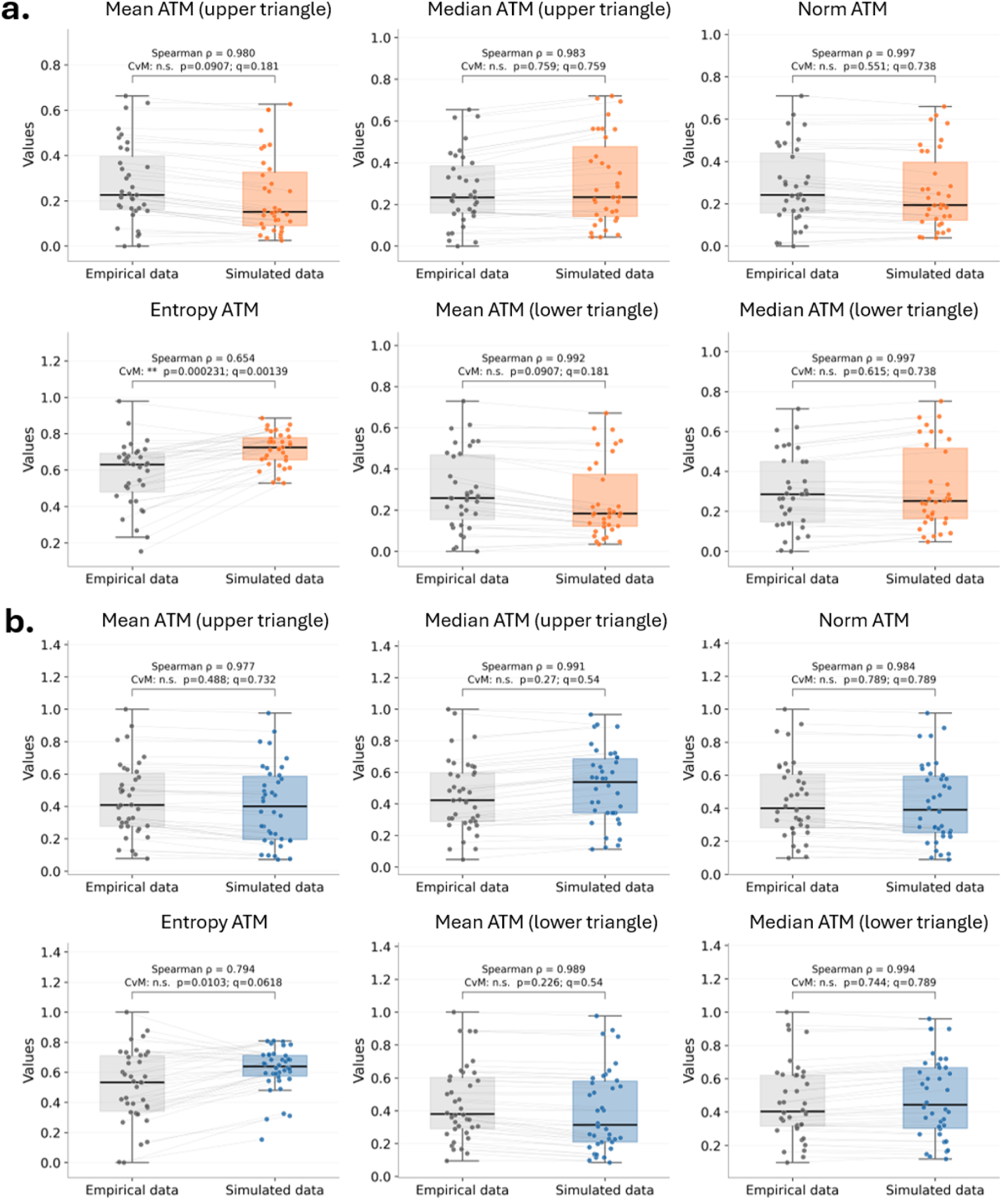
Comparison of empirical and simulated normalized avalanche transition matrix features. Empirical values (gray) and simulated predictions are shown for ALS participants (*n* = 38, blue) and controls (*n* = 35, orange). For each subject, ten stochastic simulations were generated at the fixed posterior mean estimates of *G*, *w_F_* and *w_NF_* , using different noise realizations. Each simulated point represents the median of the corresponding feature across these ten simulations. Empirical and simulated features were transformed using their respective original min-max scalers. The six panels display the mean and median of the strictly upper-triangular ATM entries, the Frobenius norm, the singular-value entropy, and the mean and median of the strictly lower-triangular entries. Boxes indicate the interquartile range, horizontal lines the median across subjects, and whiskers the most extreme observations within 1.5 interquartile ranges of the box. All individual observations are displayed, and thin lines connect empirical and simulated values from the same subject. Panel annotations report subject-level Spearman correlations (*ρ*) and the unadjusted *p*-values and FDR-adjusted *q*-values from exploratory two sample Cramér-von Mises (CvM) comparisons. FDR correction was applied across the six features separately within each group. Asterisks denote *q <* 0.05 (*), *q <* 0.01 (**), and *q <* 0.001 (***); n.s., not significant. The CvM comparisons do not account for subject pairing.

A complementary test of the simulated dynamics was obtained by examining neuronal avalanche-size and duration distributions (Supplementary Figs. SM6–SM9). Both empirical recordings and subject-specific simulations exhibited broad, decreasing distributions, indicating qualitative similarities in avalanche-like activity. The estimated avalanche-size exponents showed no statistically detectable difference between empirical and simulated data in controls after multiple-comparison correction, whereas simulated exponents were significantly higher in ALS patients. For avalanche durations, simulated exponents were systematically lower than their empirical counterparts in both groups. These discrepancies are informative because the inference procedure was constrained by ATM-derived features describing sequential regional recruitment within avalanches, whereas the distributions of avalanche size and duration were not directly fitted. Although avalanche duration may indirectly influence the number of transitions contributing to an ATM, neither the total amplitude-weighted avalanche size nor the full distribution of event durations are explicitly encoded by the selected summary statistics. The differences mentioned above identify aspects of the empirical dynamics that the present model does not fully capture and may provide useful additional constraints for future model development. Furthermore, approximate scaling over a restricted range should not be interpreted as definitive evidence of criticality or of a unique power-law generating mechanism. Several limitations should be considered when interpreting these findings. First, the reduced Wong–Wang model represents recurrent excitation without explicitly resolving inhibitory populations. The inferred parameters may therefore reflect effective changes in the excitation–inhibition balance, but the model cannot distinguish altered excitatory feedback from changes in inhibitory regulation. Second, all participants were simulated using a common structural connectome [43] derived from healthy subjects, rather than individual connectomes. Because recurrent excitation, global coupling, and network topology jointly influence whole-brain dynamics, the inferred parameters may partially accommodate effects of unmodeled individual or disease-related structural differences. Estimating global coupling as a free parameter does not eliminate this limitation, since a scalar parameter cannot reproduce changes in connection topology. Finally, independent physiological measures of cortical excitability, such as TMS-derived indices, were unavailable for this cohort. Future studies should assess sensitivity to alternative connectomes and incorporate patient-specific structural connectivity where feasible.

Overall, the present study links alterations in large-scale avalanche propagation in ALS to changes in the recurrent-excitation parameters of a whole-brain generative model. The results support a distributed increase in model-derived recurrent excitation, with a larger observed motor-frontal group effect and an association between motor-frontal excitation and functional impairment. Beyond the discrimination of patients from controls, the main value of the framework is its capacity to provide a mechanistically interpretable representation of empirical dynamics and to generate model-dependent predictions that can be tested in future physiological and longitudinal studies.

## 4 Methods

### 4.1 Whole brain modelling

In order to accurately capture the brain dynamics with a minimal model while keeping biophysical interpretability [47], the reduced Wong–Wang model was utilized. The model focuses on how local recurrent excitation, long-range coupling between regions, and background input/noise shape brain dynamics over time. The version we used, derived from [27] and described in [28] represents the global brain dynamics of a network of interconnected local units represented by the following set of coupled nonlinear stochastic differential equations:

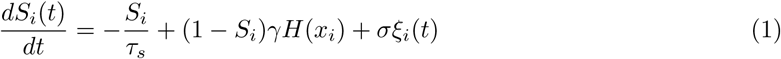

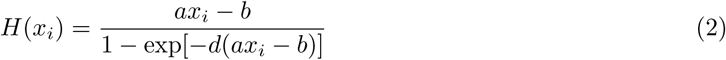

where each brain region *i* is described by the synaptic gating variable *S_i_*(*t*); *τ_s_* represents the synaptic time constant, *γ* corresponds to the kinetic parameter scaling synaptic response, and *σξ_i_*(*t*) is the noise term (Gaussian white noise). *H*(*x_i_*) is the population firing rate of the local cortical area *i* (from 1 to *N* = 80 according to the AAL Atlas) and it transforms *x_i_*, that is the total synaptic input current to each region, into the firing rate. The input to region *i* is described by the following equation:

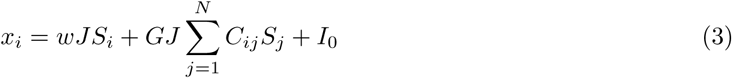

where *w* is the local recurrent excitability (self-excitation strength). More specifically, local recurrent excitability controls how strongly neurons within the same region excite each other. For example, if *w* is high, self-excitation will be stronger. This leads to more persistent and amplified activity and so, can be associated with cortical hyperexcitability. *w* reppresents our parameter of interest and is the one that will be inferred (see below). We considered two parameterizations of the recurrent excitation. In the spatially uniform model,

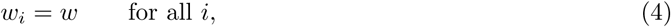

and recurrent excitation and global coupling are inferred jointly as (*w, G*). In the regional model,

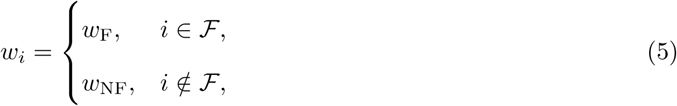

Here, F denotes the predefined motor-frontal partition, comprising 34 anatomical regions that include frontal association areas and motor-related regions, while its complement comprises the remaining 46 regions. This partition was defined anatomically, rather than on the basis of functional connectivity or empirical group differences. The complete list of regions assigned to each partition is provided in Supplementary Table 1. For this model, all three parameters, (*w*_F_*, w*_NF_*, G*), were inferred jointly from whole-brain ATM features. *J*is the synaptic coupling constant, G is the global coupling strength between regions, *C_ij_* is the structural connectivity weight from region *j* to *i* expressing the neuroanatomical links between cortical region *i* and *j* ; *I*_0_ is the external background input. In particular, the first term represents the local recurrent excitation, the second term the long-range input from the other regions. Parameter values for the input– output function were set as *a* = 250(*n/C*), *b* = 160(*Hz*), and *d* = 0.05(*s*). The kinetic parameters are *γ* = 0.641*/*1000, and *τ_s_* = 100*ms*. The synaptic couplings are *J* = 0.2609(*nA*), and the overall effective external input is *I*_0_ = 0.382(*nA*). Global coupling *G* was estimated for each participant. The inference procedure retained posterior dependencies among the three parameters. Summarizing the dynamics governing the system, at each moment a region receives its own recurrent excitation, the inputs from the connected regions, and background drive. These inputs modulate the firing rate through the nonlinear function *H*(*x_i_*) which updates synaptic activity *S_i_*, as explained above. Within this framework, increased local recurrent excitability (*w*) represents cortical hyperexcitability observed in ALS. All simulations were performed using the same group-averaged structural connectome derived from 47 healthy participants, as described in Section 4.3. The structural connectivity matrix was held fixed across subjects and model configurations, whereas recurrent excitation and global coupling were varied or inferred according to the considered analysis. Therefore, the estimated parameters should be interpreted as conditional on this common anatomical scaffold. Within this model, increasing *w* strengthens recurrent excitatory feedback within a region, whereas increasing *G* strengthens input transmitted through the structural network. The inferred excitation parameters thus provide model-derived measures of recurrent excitation whose elevation is compatible with cortical hyperexcitability, rather than constituting direct physiological measurements of excitability.

### Numerical simulations

The stochastic differential equations were integrated using the Heun scheme with a time step of Δ*t* = 1 ms. Independent Gaussian noise increments were generated for each brain region and integration step as 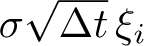, where *ξ_i_* ∼ N (0, 1) and *σ* = 0.05, with time expressed in milliseconds. Each simulation lasted 300 s, with the first 60 s discarded as a transient. The same numerical protocol was used throughout each simulation ensemble.

### 4.2 Participants and data acquisition

A total of 39 patients diagnosed with Amyotrophic Lateral Sclerosis (ALS) were included in the study (29 males, 10 females; mean age 59.63±12.87 years; mean education 10.38±4.3 years). Patients were recruited from the Hermitage Capodimonte Clinic in Naples and affiliated centers. Diagnosis was established according to the revised El Escorial criteria as described in [20]. Inclusion criteria required the absence of major medical conditions or medications that could affect MEG recordings, as well as the absence of other significant neurological, psychiatric, or systemic disorders. Structural MRI scans were screened to exclude focal or diffuse brain abnormalities. The control group consisted of 39 healthy individuals (28 males, 11 females), matched for age (64.0 ± 10.4 years) and education (12.0 ± 4.3 years) [48]. Functional status in ALS patients was assessed using the ALS Functional Rating Scale-Revised (ALSFRS-R) [12], where higher scores indicate better functional capacity. Disease staging was further evaluated using the Milano–Torino Staging (MiToS) system [49], derived from ALSFRS-R subscores. The study protocol was approved by the Local Ethics Committee of the University of Campania “Luigi Vanvitelli” (Italy), and all participants provided written informed consent in accordance with the Declaration of Helsinki.

#### MEG acquisition and preprocessing

Resting-state MEG data were acquired in a magnetically shielded room using a system equipped with 154 SQUID magnetometers and 9 reference sensors. Head position was continuously monitored using a Polhemus Fastrack digitizer, based on four anatomical landmarks and four head position indicator coils.

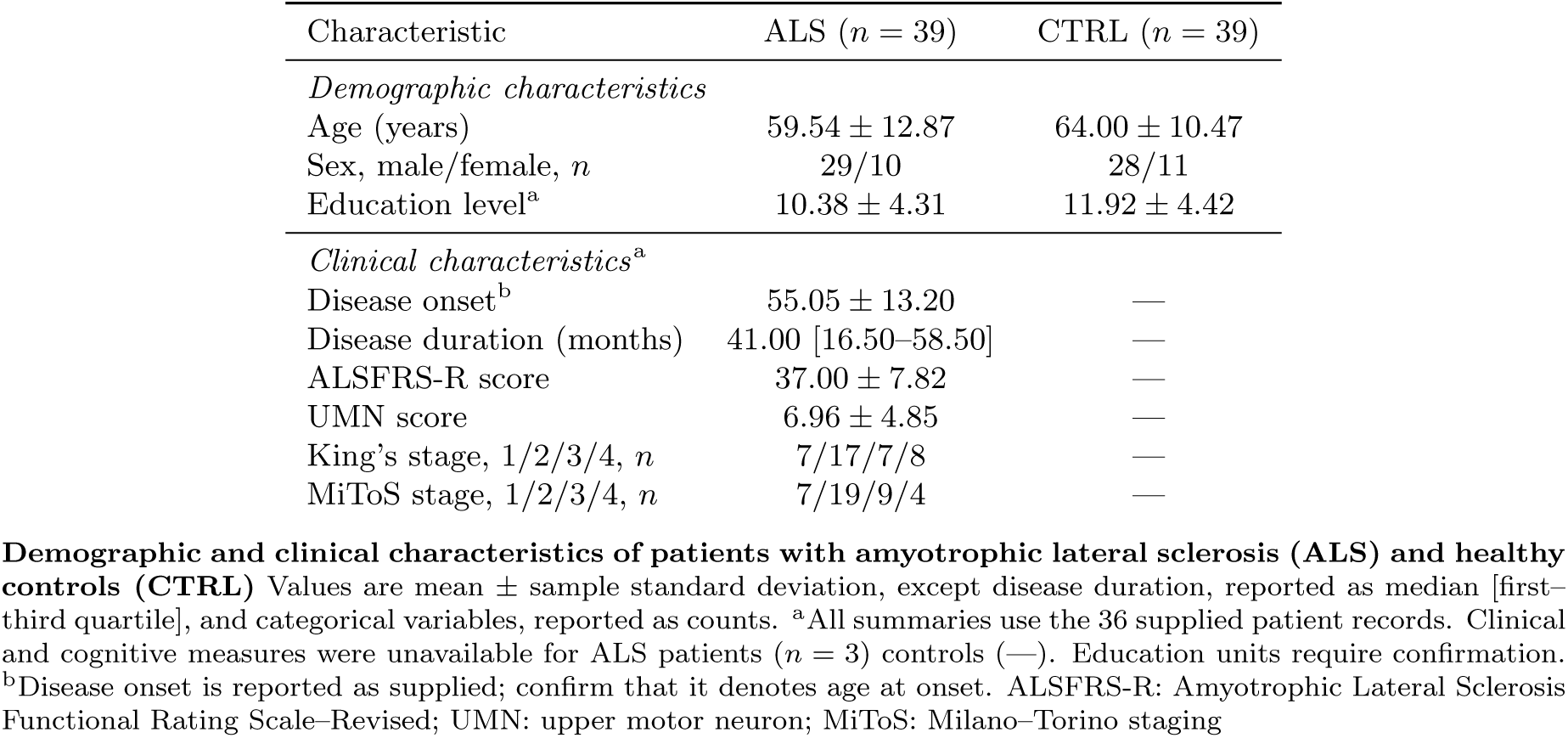

| Characteristic | ALS (*n* = 39) | CTRL (*n* = 39) |
|---|---|---|
| *Demographic characteristics* |  |  |
| Age (years) | 59.54 ± 12.87 | 64.00 ± 10.47 |
| Sex, male/female, *n* | 29/10 | 28/11 |
| Education level<sup>a</sup> | 10.38 ± 4.31 | 11.92 ± 4.42 |
| *Clinical characteristics*<sup>a</sup> |  |  |
| Disease onset<sup>b</sup> | 55.05 ± 13.20 | — |
| Disease duration (months) | 41.00 [16.50–58.50] | — |
| ALSFRS-R score | 37.00 ± 7.82 | — |
| UMN score | 6.96 ± 4.85 | — |
| King’s stage, 1/2/3/4, *n* | 7/17/7/8 | — |
| MiToS stage, 1/2/3/4, *n* | 7/19/9/4 | — |

Values are mean ± sample standard deviation, except disease duration, reported as median [first–third quartile], and categorical variables, reported as counts.
<sup>a</sup>All summaries use the 36 supplied patient records. Clinical and cognitive measures were unavailable for ALS patients (*n* = 3) controls (—). Education units require confirmation.
<sup>b</sup>Disease onset is reported as supplied; confirm that it denotes age at onset. ALSFRS-R: Amyotrophic Lateral Sclerosis Functional Rating Scale–Revised; UMN: upper motor neuron; MiToS: Milano–Torino staging

Each participant underwent two resting-state recordings (3.5 minutes each) with eyes closed, separated by a one-minute pause. Electrocardiogram (ECG) and electrooculogram (EOG) signals were recorded simultaneously to facilitate artifact identification. Data were anti-alias filtered and sampled at 1024 Hz.

Preprocessing was performed using the FieldTrip toolbox in MATLAB. Signals were band-pass filtered between 0.5 and 48 Hz using a fourth-order Butterworth filter. Environmental noise was reduced via principal component analysis (PCA), and physiological artifacts were removed using independent component analysis (ICA), typically identifying one cardiac component and, less frequently, ocular components. We obtained recordings during the eyes closed condition, which drastically reduces blink-related activity. After excluding segments contaminated by excessive noise, the remaining data from both runs were concatenated to produce a cleaned time series of equal length for each participant (122.8 seconds).

#### Source-reconstruction data

MEG source reconstruction was performed using the FieldTrip toolbox [50] followed the methods outlined by [21]. For each participant, MEG recordings were co-registered with their native structural MRI using four anatomical fiducial points digitized before acquisition: the nasion, the left and right preauricular points, and the vertex. A subject-specific single-shell volume conduction model was constructed according to Nolte (2003) to solve the forward problem. Source-level neural activity was subsequently estimated using a linearly constrained minimum variance (LCMV) beamformer ([51, 52]), which spatially filters sensor-level recordings to estimate activity at predefined source locations. The source space was parcellated according to the Automated Anatomical Labeling (AAL) atlas ([53]), and regional time series were reconstructed at the centroid of each region of interest (ROI). Of the original 116 AAL regions, we excluded the 26 cerebellar regions and 10 non-hippocampal subcortical regions, retaining the bilateral hippocampi and obtaining 80 source regions., resulting in 80 regional source-level time series per participant. We visually inspected the reconstructed signals to identify and exclude segments containing residual artifacts before subsequent analyses. Subcortical and cerebellar regions were excluded due to, resulting in a final set of *N* = 80 cortical regions per subject. The list of regions is reported in Table 1.

**Table 1:** Regional partition of the 80 AAL atlas regions included in the model. Regions were assigned to a motor-frontal partition (34 regions), governed by the regional excitability parameter *w_F_* , or to the remaining-region partition (46 regions), governed by *w_NF_* . The motor-frontal partition comprises the frontal and motor-related regions listed below. The suffixes L and R indicate the left and right hemispheres, respectively.

| Index | Region | Model partition (Motor-Frontal/Remaining regions) |
|---|---|---|
| 0 | Rectus_L | Motor-Frontal |
| 1 | Olfactory_L | Motor-Frontal |
| 2 | Frontal_Sup_Orb_L | Motor-Frontal |
| 3 | Frontal_Med_Orb_L | Motor-Frontal |
| 4 | Frontal_Mid_Orb_L | Motor-Frontal |
| 5 | Frontal_Inf_Orb_L | Motor-Frontal |
| 6 | Frontal_Sup_L | Motor-Frontal |
| 7 | Frontal_Mid_L | Motor-Frontal |
| 8 | Frontal_Inf_Oper_L | Motor-Frontal |
| 9 | Frontal_Inf_Tri_L | Motor-Frontal |
| 10 | Frontal_Sup_Medial_L | Motor-Frontal |
| 11 | Supp_Motor_Area_L | Motor-Frontal |
| 12 | Paracentral_Lobule_L | Motor-Frontal |
| 13 | Precentral_L | Motor-Frontal |
| 14 | Rolandic_Oper_L | Motor-Frontal |
| 15 | Postcentral_L | Remaining regions |
| 16 | Parietal_Sup_L | Remaining regions |
| 17 | Parietal_Inf_L | Remaining regions |
| 18 | SupraMarginal_L | Remaining regions |
| 19 | Angular_L | Remaining regions |
| 20 | Precuneus_L | Remaining regions |
| 21 | Occipital_Sup_L | Remaining regions |
| 22 | Occipital_Mid_L | Remaining regions |
| 23 | Occipital_Inf_L | Remaining regions |
| 24 | Calcarine_L | Remaining regions |
| 25 | Cuneus_L | Remaining regions |
| 26 | Lingual_L | Remaining regions |
| 27 | Fusiform_L | Remaining regions |
| 28 | Heschl_L | Remaining regions |
| 29 | Temporal_Sup_L | Remaining regions |
| 30 | Temporal_Mid_L | Remaining regions |
| 31 | Temporal_Inf_L | Remaining regions |
| 32 | Temporal_Pole_Sup_L | Remaining regions |
| 33 | Temporal_Pole_Mid_L | Remaining regions |
| 34 | ParaHippocampal_L | Remaining regions |
| 35 | Cingulum_Ant_L | Motor-Frontal |
| 36 | Cingulum_Mid_L | Motor-Frontal |
| 37 | Cingulum_Post_L | Remaining regions |
| 38 | Insula_L | Remaining regions |
| 39 | Rectus_R | Motor-Frontal |
| 40 | Olfactory_R | Motor-Frontal |
| 41 | Frontal_Sup_Orb_R | Motor-Frontal |
| 42 | Frontal_Med_Orb_R | Motor-Frontal |
| 43 | Frontal_Mid_Orb_R | Motor-Frontal |
| 44 | Frontal_Inf_Orb_R | Motor-Frontal |
| 45 | Frontal_Sup_R | Motor-Frontal |
| 46 | Frontal_Mid_R | Motor-Frontal |
| 47 | Frontal_Inf_Oper_R | Motor-Frontal |
| 48 | Frontal_Inf_Tri_R | Motor-Frontal |
| 49 | Frontal_Sup_Medial_R | Motor-Frontal |
| 50 | Supp_Motor_Area_R | Motor-Frontal |
| 51 | Paracentral_Lobule_R | Motor-Frontal |
| 52 | Precentral_R | Motor-Frontal |
| 53 | Rolandic_Oper_R | Motor-Frontal |
| 54 | Postcentral_R | Remaining regions |
| 55 | Parietal_Sup_R | Remaining regions |
| 56 | Parietal_Inf_R | Remaining regions |
| 57 | SupraMarginal_R | Remaining regions |
| 58 | Angular_R | Remaining regions |
| 59 | Precuneus_R | Remaining regions |
| 60 | Occipital_Sup_R | Remaining regions |
| 61 | Occipital_Mid_R | Remaining regions |
| 62 | Occipital_Inf_R | Remaining regions |
| 63 | Calcarine_R | Remaining regions |
| 64 | Cuneus_R | Remaining regions |
| 65 | Lingual_R | Remaining regions |
| 66 | Fusiform_R | Remaining regions |
| 67 | Heschl_R | Remaining regions |
| 68 | Temporal_Sup_R | Remaining regions |
| 69 | Temporal_Mid_R | Remaining regions |
| 70 | Temporal_Inf_R | Remaining regions |
| 71 | Temporal_Pole_Sup_R | Remaining regions |
| 72 | Temporal_Pole_Mid_R | Remaining regions |
| 73 | ParaHippocampal_R | Remaining regions |
| 74 | Cingulum_Ant_R | Motor-Frontal |
| 75 | Cingulum_Mid_R | Motor-Frontal |
| 76 | Cingulum_Post_R | Remaining regions |
| 77 | Insula_R | Remaining regions |
| 78 | Hippocampus_L | Remaining regions |
| 79 | Hippocampus_R | Remaining regions |

### 4.3 Tractography data

The structural connectivity matrix used in this study was obtained from the healthy cohort previously investigated by Sorrentino et al. [43]. Specifically, we employed a group-averaged structural connectome derived from diffusion MRI tractography data acquired from 47 healthy participants. The reconstruction procedure, including diffusion MRI preprocessing, deterministic tractography, and estimation of interregional connectivity weights [43]. High-resolution 3D T1-weighted brain images were acquired on a 1.5 T Signa system (GE Healthcare) using a 3D magnetization-prepared gradient-echo (BRAVO) sequence (TR/TE/TI = 8.2/3.1/450 ms; isotropic voxel size = 1 × 1 × 1 mm^3^; 50% partition overlap; 324 sagittal slices providing whole-brain coverage). Diffusion MRI data for subject-specific connectome reconstruction were collected using echo-planar imaging (TR/TE = 12,000/95.5 ms; voxel size = 0.94 × 0.94 × 2.5 mm^3^), with 32 diffusion-weighted directions and 5 non-diffusion-weighted (B0) volumes. MRI acquisition followed MEG recording. Diffusion data preprocessing was performed using tools from the FMRIB Software Library (FSL, http://fsl.fmrib.ox.ac.uk/fsl) [43]. All datasets were corrected for head motion and eddy current-induced distortions using the eddy_correct routine, with appropriate rotation of diffusion gradient directions. Brain masks were generated from B0 images using the Brain Extraction Tool. A diffusion tensor model was fitted voxel-wise, and whole-brain streamline reconstruction was carried out via deterministic tractography using Diffusion Toolkit, implementing the FACT propagation algorithm (angle threshold = 45*^◦^*, spline interpolation, and masking based on fractional anisotropy maps thresholded at 0.2). For connectivity analysis, regions of interest (ROIs) were defined according to the Automated Anatomical Labeling (AAL) atlas in MNI space. The atlas was constrained by gray matter tissue probability maps derived from Statistical Parametric Mapping (SPM; threshold = 0.2). Individual FA maps were spatially normalized to MNI space using the FSL FA template and the normalization routine implemented in SPM12. The resulting transformation matrices were inverted and applied to warp ROIs into each subject’s native space. Normalization quality was verified through visual inspection. Structural connectivity was represented by a matrix *C_ij_*, whose entries quantify the strength of anatomical connections between brain regions. The connectome was aligned with the 80 regions considered in the present whole-brain model. The same group-averaged structural connectivity matrix was used for all simulations and was held fixed across ALS patients and healthy controls. This common anatomical scaffold was employed in both the spatially uniform and regional model configurations, ensuring that differences in simulated dynamics arose from variations in the model parameters rather than from differences in the underlying structural network. Consequently, individual variability and ALS-related alterations in structural connectivity were not explicitly represented in the model.

### 4.4 Feature Extraction

We characterized whole-brain dynamics using six summary features derived from directed Avalanche Transition Matrices (ATMs): the mean and median of the strictly upper and lower triangles, the Frobenius norm, and singular-value-based entropy. The same feature definitions and regional ordering were used for empirical recordings and simulated activity.

#### Neuronal avalanche detection and Avalanche Transition Matrices

Starting from source-reconstructed MEG signals, each regional time series is first standardized using a z-score transformation. To determine the activation threshold, we examined the distribution of standardized activity across participants and identified the point at which it deviated from a Gaussian distribution [42]. This deviation occurred at approximately 2.5 standard deviations, which was therefore adopted as the threshold for identifying suprathreshold activity. Time points exceeding this threshold are considered active, resulting in a binarized representation of the data. Based on this representation, neuronal avalanches are defined as contiguous periods of activity bounded by time bins in which no regions are active. The size of an avalanche is defined as the cumulative suprathreshold amplitude excess of the z-scored regional signals, summed over all regions and time samples belonging to each avalanche. Specifically, for an activation threshold of (|*σ*| = 2.5), avalanche size was calculated as ∑*_r,t∈A_* max(|*z_r_*(*t*)|−2.5, 0). Avalanche duration corresponds to the number of consecutive time bins during which the avalanche persists. To validate the presence of neuronal avalanches, we examine the distributions of avalanche sizes and durations, which are reported in the Supplementary Material. These distributions exhibit power-law behavior, consistent with the characteristic signatures of avalanche dynamics in neural systems [54, 55].

Within each detected avalanche, we analyze the spatiotemporal propagation of activity by constructing avalanche transition matrices (ATMs) [20]. Each ATM encodes the probability that activation in one region is followed by activation in another region in the subsequent time step. Avalanches shorter than two samples were excluded. Avalanche-specific transition estimates were aggregated into a subject-level Avalanche Transition Matrix (ATM). The matrices were retained in their directed, non-symmetrized form, preserving differences between opposite transition directions. Accordingly, ATM*_ij_*and ATM*_ji_* were not constrained to be equal. Directionality denotes temporal ordering of suprathreshold activations and does not establish causal influence.

Subsequently, data features are extracted from the avalanche transition matrices (ATMs), including the mean value, median for the upper and lower triangles to take into account the directionality of the transitions, Frobenius norm and entropy. For each participant, the mean ATM was calculated as the arithmetic mean of the entries included in the analyses. It summarizes the average transition probability across regional pairs and therefore characterizes the average propensity for avalanche-related activity propagation. The Frobenius norm was calculated as the square root of the sum of squared matrix entries and quantifies the overall magnitude of the transition weights. Unlike the mean, it gives greater weight to larger entries. To characterize the organization of the directed matrix, we computed Shannon entropy from its normalized singular values. For the singular value decomposition *A* = *U* Σ*V* ^T^, let *s_k_* denote the singular values and define

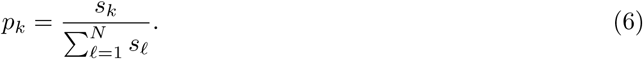

ATM entropy was then calculated as

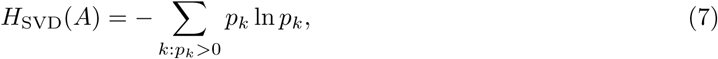

using the full matrix, including its diagonal. Natural logarithms were used, yielding entropy in nats. Entropy was undefined for an all-zero matrix. Unlike an eigenvalue-based construction, the singular values are nonnegative and well-defined for an asymmetric ATM. Higher entropy indicates a more even distribution of singular-value weight across components; lower entropy indicates that fewer components dominate the matrix structure. This measure does not directly quantify the number of distinct avalanche patterns.

#### Empirical statistical comparisons

For each metric shared between the ALS and CTRL groups, potential outliers were identified separately within each group using the interquartile range (IQR) criterion, defined as values lying outside the interval [*Q*1 − 1.5 · *IQR, Q*3 + 1.5 · *IQR*]. Outlier observations were excluded from both statistical analyses and clinical correlations. Group differences were then assessed on the resulting datasets using Welch’s two-sample t-test, which does not assume equal variances between groups.

For the model inversion procedure, all selected metrics were subsequently normalized to the [0,1] range using min–max scaling. This was done to place heterogeneous features on a common scale and to ensure that the inference procedure focused on reproducing the relative pattern of alterations observed across empirical metrics, rather than the absolute values of the individual metrics. Accordingly, the objective of the inversion was not to match the exact magnitude of each individual metric, but to capture the overall trend and relationships characterizing the empirical data.

#### Clinical associations

Associations between subject-level parameter estimates and ALSFRS-R, King’s stage, and MiToS stage were assessed using two-sided Spearman rank-order correlations. Each participant contributed one posterior mean per parameter; posterior samples were not treated as independent observations. Analyses used participants with available values for the parameter and clinical outcome. Correlations were additionally examined in the subgroup whose posterior means satisfied *w̄*_F_ *> w̄*_NF_. This subgroup analysis was exploratory. Statistical significance after FDR correction was defined as an adjusted *p*-value below 0.05.

### 4.5 Bayesian model inversion via simulation-based inference and pipeline validation

The Bayesian framework provides a principled approach for parameter estimation, prediction, and uncertainty quantification by updating prior knowledge with evidence from empirical observations [56]. In this setting, uncertainty is explicitly represented through probability distributions: prior beliefs about model parameters are updated using observed data to obtain posterior distributions. However, performing Bayesian inference on large-scale brain data is challenging due to the high dimensionality of the observations, the nonlinearity of brain dynamics, and the complex interactions imposed by the underlying brain network structure. In particular, the likelihood function is typically computationally intractable for such models, making conventional approaches such as Markov Chain Monte Carlo (MCMC) computationally expensive [39].

To overcome these limitations, we adopt a likelihood-free approach based on Simulation-Based Inference (SBI) [57, 58], in which the computational model is treated as a stochastic simulator capable of generating synthetic data that resemble empirical observations. This framework enables parameter inference without explicitly evaluating the likelihood function. Specifically, we use neural density estimation techniques based on Normalizing Flows (NFs), which learn flexible transformations from simple base distributions (e.g., the prior) to complex posterior distributions. By construction, these transformations are invertible and differentiable, enabling efficient sampling and exact likelihood or density evaluation.

Our objective is to infer the posterior distribution of the model excitability parameters that best explain the observed empirical features. To this end, we considered two complementary model configurations. In the first spatially uniform configuration, a single global excitability parameter (*w*) controlled the excitability of all brain regions, while *G* scaled interregional coupling. Joint inference of (*w, G*) addressed whether the empirical dynamics were compatible with increased recurrent excitation when global coupling was also allowed to vary. This configuration therefore provided a parsimonious assessment of a spatially uniform alteration in excitation without fixing the coupling regime. In the partition-specific configuration, the uniform excitation parameter was replaced by *w*_F_ for the 34 motor-frontal regions and *w*_NF_ for the remaining 46 regions. Global coupling *G* remained an inferred parameter. Joint inference of (*w*_F_*, w*_NF_*, G*) allowed us to examine whether the empirical dynamics were compatible with different excitation levels across the two anatomical partitions. The uniform model is recovered when *w*_F_ = *w*_NF_. The two configurations thus address complementary questions concerning the overall elevation and regional distribution of recurrent excitation. They were fitted separately; estimates from the uniform configuration were not used to fix *G* in the partition-specific inference. Independent uniform priors were assigned to the parameters. Recurrent-excitation parameters were sampled over [2, 4], and global coupling over [1.5, 2.5]:

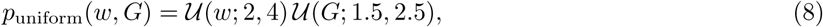

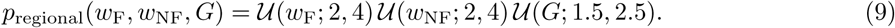

Note that the independence of the priors does not impose independence on the inferred posterior distributions. We employ Sequential Neural Posterior Estimation (SNPE) [58], a simulation-based inference (SBI) framework that learns the relationship between model parameters and observed summary statistics directly from simulations. This approach is particularly well-suited for mechanistic whole-brain models, for which the likelihood functions are generally intractable. The posterior density is approximated using a Masked Autoregressive Flow (MAF) [59], a class of normalizing flows that enables flexible and computationally efficient posterior density estimation through a sequence of invertible nonlinear transformations. SNPE requires three key components: (i) a prior distribution over parameters, (ii) a generative model capable of simulating brain activity for a given parameter configuration, and (iii) a set of low-dimensional features summarizing the empirical data. In our case, the generative model is the reduced Wong-Wang whole-brain neural mass model, and the features are computed from source-reconstructed signals. Specifically, we extract a set of summary statistics, as described in the previous paragraph.

For participant *s*, the trained estimators were conditioned on the empirical feature vector **y***_s,_*_expt_ to approximate

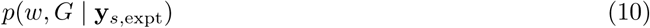

for the uniform model configuration, and

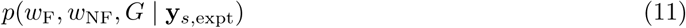

for the partition-specific model configuration. Both configurations used the same six whole-brain ATM features computed on the whole brain. Regional excitation parameters were therefore inferred from their combined effects on whole-brain dynamics, rather than from features computed separately within each anatomical partition. Joint posterior sampling retained dependencies among excitation and coupling parameters, including potential compensatory relationships. We generated 4,000 simulations for the spatially uniform configuration, jointly inferring (*w, G*), and 20,000 simulations for the partition-specific configuration, jointly inferring (*w*_F_*, w*_NF_*, G*). The parameter vectors were sampled from the corresponding prior distributions. Held-out simulations used for synthetic validation were excluded from training. Once trained, each conditional density estimator was applied to participants’ empirical feature vectors to evaluate and sample from the approximate posterior without additional simulations. This amortized inference procedure does not require the generative model to be differentiable.

Before applying empirical observations, we evaluated parameter recovery using held-out synthetic simulations with known ground-truth parameters. Test simulations were excluded from network training and generated using the same structural-connectivity matrix and simulation protocol as the training data. The uniform-model ensemble comprised 4,000 simulations, of which 3,200 were assigned to training and 800 to held-out testing. The regional-model ensemble comprised 20,000 simulations, partitioned into 16,000 training and 4,000 test simulations. For each test simulation, the trained estimator was conditioned on the corresponding feature vector, and joint posterior samples were drawn. Parameter recovery was evaluated by comparing posterior means with ground-truth values. Marginal posterior uncertainty was summarized using posterior standard deviations and equal-tailed 95% credible intervals. The quality of the inferred posteriors is assessed using diagnostic measures including posterior shrinkage and z-scores, defined as *z* = (*θ*^^^− *θ*_true_)*/σ*_posterior_, where *θ*^^^ is the posterior mean, *θ*_true_ is the ground-truth parameter, and *σ*_posterior_ is the posterior standard deviation.

We additionally evaluated marginal empirical coverage as the proportion of test simulations for which the 95% credible interval contained the ground-truth parameter. Coverage was compared with the nominal 95% level, and recovery plots were examined across the parameter range to identify regions with larger errors. High shrinkage was interpreted as posterior concentration rather than evidence of accurate recovery or calibration. The illustrated posterior examples complement the evaluation across the full held-out set. The resulting estimates exhibit low z-scores and high shrinkage, indicating accurate and informative Bayesian inference across the explored parameter range. Parameter-dependent recovery analyses further revealed localized biases and reduced coverage near portions of the prior boundaries. (see Fig. 5d,g, Fig. 7, c,f,i). To run SNPE, we used the public SBI toolbox [60].

#### Synthetic parameter recovery

For the first inference pipeline. Parameter recovery was evaluated on *N* = 800 held-out synthetic simulations. For each test case *i*, let *θ_i_* denote the true generating parameter and *θ*^^^*_i_* the corresponding posterior mean. Recovery accuracy was quantified using the mean absolute error (MAE), root mean squared error (RMSE), and signed bias:

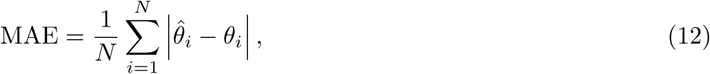

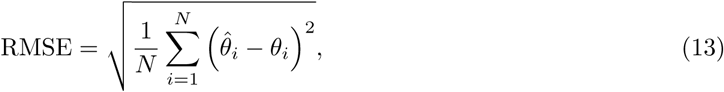

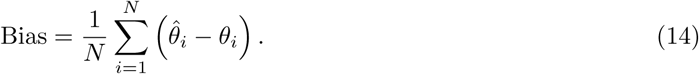

Positive and negative bias indicate systematic overestimation and underestimation, respectively. Errors were calculated in the original parameter units. For each test case, the equal-tailed 95% credible interval was defined by the 2.5th and 97.5th percentiles of the posterior samples, [*q*_0.025*,i*_*, q*_0.975*,i*_]. Empirical coverage was calculated as

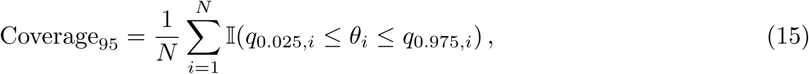

where I(·) is the indicator function. Coverage was reported as a percentage. The test simulations were excluded from neural-network training. The values of this analysis are reported in table 2 in the Supplementary Material.

**Table 2:** Synthetic parameter recovery for the global and regional models. The global model was jointly inferred *w* and *G* using a 4,000-simulation ensemble, with 800 held-out test cases. The regional model jointly inferred *w_F_* , *w_NF_* , and *G* using a 20,000-simulation ensemble, with 4,000 held-out test cases. Both models used six normalized ATM features. Point estimates were posterior means, and errors were calculated in the original parameter units. Bias denotes the posterior mean minus the true generating value. Coverage is the percentage of true values contained within the equal-tailed 95% credible intervals. Test cases were excluded from neural-network training.

| Model | Parameter | *N*<sub>test</sub> | MAE | RMSE | Bias | Coverage 95% |
|---|---|---|---|---|---|---|
| Global | *w* | 800 | 0.062682 | 0.085447 | +0.005641 | 93.125% |
| Global | *G* | 800 | 0.114182 | 0.143605 | +0.000049 | 91.375% |
| Regional | *w*<sub>*F*</sub> | 4000 | 0.095805 | 0.128848 | +0.006991 | 92.775% |
| Regional | *w*<sub>*NF*</sub> | 4000 | 0.092195 | 0.124301 | −0.013293 | 92.650% |
| Regional | *G* | 4000 | 0.110675 | 0.140004 | +0.012295 | 92.450% |

#### Reproduction of empirical features from inferred parameters

For each of the 73 non-outlier subjects (38 ALS and 35 controls), the posterior means of *w_F_* , *w_NF_* , and *G* were used to generate ten stochastic simulations. The parameter triplet was held fixed across simulations, while the random seed was varied to obtain distinct noise realizations. All simulations followed the numerical protocol described above, and the same six ATM features were extracted from each realization. For each subject and feature, the median across the ten simulations was used as the simulated prediction. These predictions were transformed using the original simulation min-max scaler, without refitting. Empirical features were transformed using the original empirical scaler, shared across ALS and controls. Comparisons were therefore performed in the normalized representation used for inference. Empirical and simulated values were displayed as adjacent box-and-whisker plots for each feature and diagnostic group. Each simulated point represented one subject’s median across ten realizations; thin lines connected empirical and simulated values from the same subject. Boxes represented the interquartile range, the central line indicated the median across subjects, and whiskers extended to the most extreme observations within 1.5 interquartile ranges of the box boundaries. Subject-level correspondence was quantified using two-sided Spearman correlations. Differences between empirical and simulated marginal distributions were assessed using the two-sample Cramér–von Mises test. Benjamini–Hochberg correction was applied across the six features within each diagnostic group, separately for correlation and distributional tests. Subjects, rather than individual simulation realizations, were the units of statistical analysis. Repeated simulations characterized stochastic variability at fixed posterior mean parameters; they did not propagate parameter uncertainty.

#### Discrimination between ALS patients and controls

We evaluated whether the inferred parameters discriminated ALS patients from healthy controls using separate classification models for each parameter. Each participant contributed one observation, represented by the corresponding posterior mean; posterior samples were not treated as independent observations. ALS was designated as the positive class. Classification was performed using L2-regularized logistic regression and nested stratified five-fold cross-validation, repeated five times with different random partitions. Identical outer splits were used across parameter-based classifiers. Within each outer training set, the inverse regularization strength was selected from *C* ∈ {0.01, 0.1, 1, 10, 100} using an inner stratified three-fold cross-validation, maximizing the mean validation ROC AUC. Predictors were standardized using statistics estimated exclusively from the corresponding training folds. For each repetition, predicted probabilities for the held-out participants were pooled across the five outer folds, yielding one out-of-fold prediction per participant. Receiver operating characteristic (ROC) curves were constructed by varying the classification threshold and plotting sensitivity against the false-positive rate. Discrimination was quantified by the area under the ROC curve (AUC). We report the mean and standard deviation of the five repetition-specific AUCs. For visualization, ROC curves were interpolated onto a common false-positive-rate grid and averaged across repetitions. This analysis assessed discrimination conditional on the previously estimated model parameters and the upstream preprocessing and subject-selection procedures. The inference pipeline was not re-estimated within each classification fold; the evaluation therefore concerns the downstream classifier rather than an independently validated end-to-end diagnostic pipeline.

## Acknowledgments

Helpful discussions with Silvia Scarpetta and Antonio de Candia are gratefully acknowledged. This work was supported by Governo Italiano Ministero per lo sviluppo Economico, ACCORDI PER INNOVAZIONE. Approccio User-friendly integrato per Diagnosi, Assistenza e Cura Efficaci—AUDACE grant number B69J23006050007.

## Data availability statement

The datasets analysed in this study are available from the corresponding author on reasonable request.

## Code availability

The code is available here: https://github.com/marangiol/Towards-the-Virtual-ALS-Patient.git

## Declaration of generative AI and AI-assisted technologies in the manuscript preparation process

During the preparation of this work, the authors used ChatGPT-Codex in order to assist with language editing and code support. After using this tool, the authors reviewed and edited the content as needed and take full responsibility for the content of the published article.

## Author Contribution

Conceptualization: M.A., P.S., D.D., and M.H. Methodology: M.A., P.S., D.D., and M.H. Investigation: P.S., D.D. Supervision: P.S., D.D. and M.H. Data collection and curation: M.D., F.T., C.G., D.T., M.D.L., E.G., G.S., P.S. Resources: D.T. and V.J. Data processing: M.D., F.T., C.G., D.T., M.D.L., E.G., G.S., P.S. Writing—original draft: M.A., P.S., D.D. and M.H. Writing—review and editing: M.A., P.S., D.D., M.D., F.T., C.G., D.T., M.D.L., E.G., G.S., V.J.

## Supplementary Materials

**Figure SM1:**
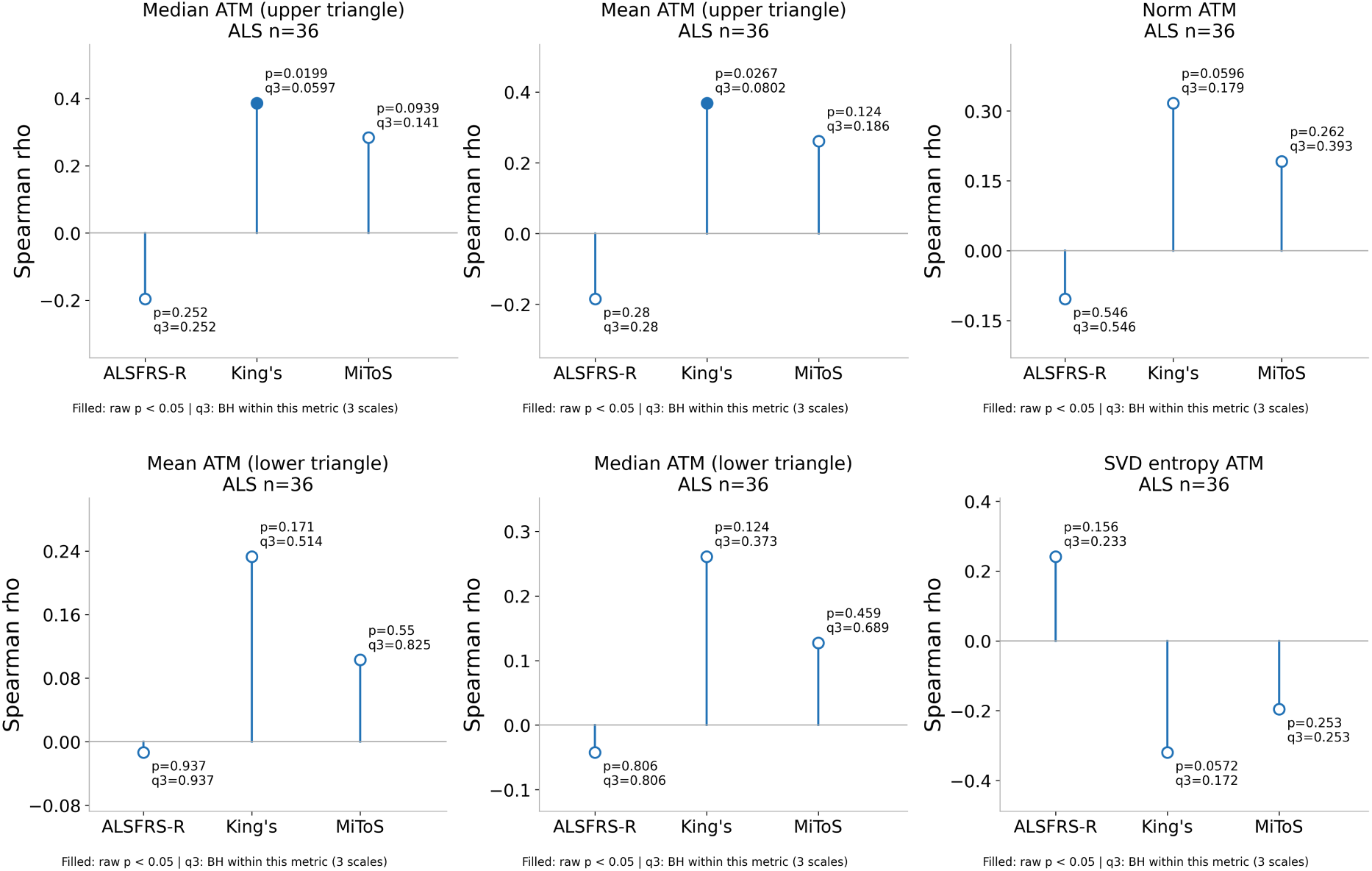
Associations between empirical avalanche transition matrix features and clinical measures in ALS. Spearman rank-order correlations between the six empirical whole-brain avalanche transition matrix (ATM) features and clinical outcomes in ALS patients. The features comprise the mean and median transition probabilities of the strictly upper and lower ATM triangles, the Frobenius norm, and the singular-value-based ATM entropy. For each feature, correlations were evaluated with the Amyotrophic Lateral Sclerosis Functional Rating Scale–Revised (ALSFRS-R), King’s clinical stage, and Milano–Torino Staging (MiToS) system. Blue stems indicate the direction and magnitude of Spearman’s correlation coefficient (*ρ*), with the horizontal gray line representing zero correlation. Although some nominal associations were observed, none survived the specified false discovery rate (FDR) correction for multiple comparisons.

**Figure SM2:**
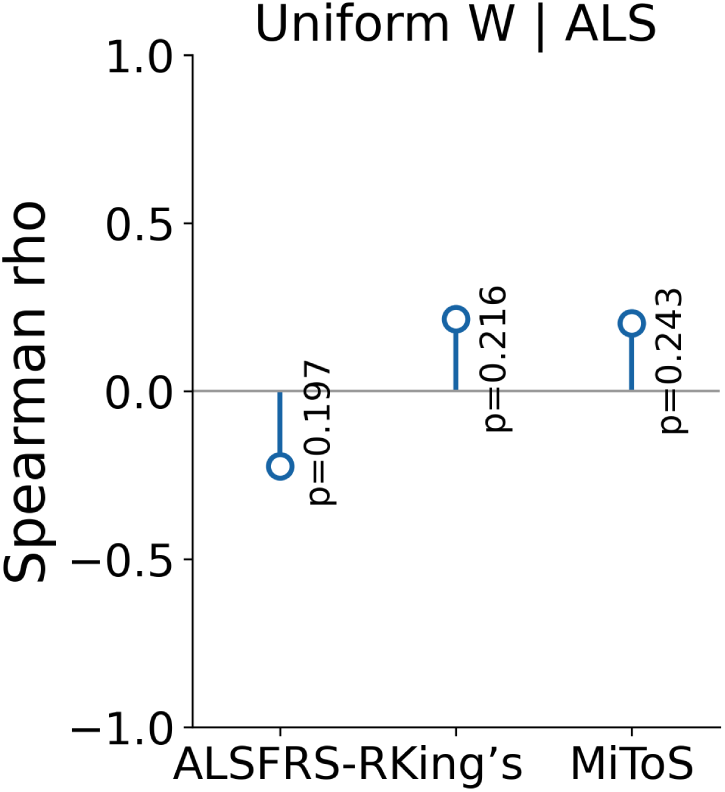
Associations between global excitability and clinical scores in ALS. Spearman correlations between individual posterior means of the spatially uniform excitability parameter *w* and ALSFRS-R, King’s stage, and MiToS stage were evaluated in 35 ALS participants, excluding the metric outlier SLA 21 and three patients with missing clinical outcome data. Stems indicate the direction and magnitude of Spearman’s *ρ*; annotations report unadjusted two-sided *p*-values. Open markers indicate associations that did not survive Benjamini-Hochberg false discovery rate correction across the three clinical scales (*q* ≥ 0.05). No association was statistically significant.

**Figure SM3:**
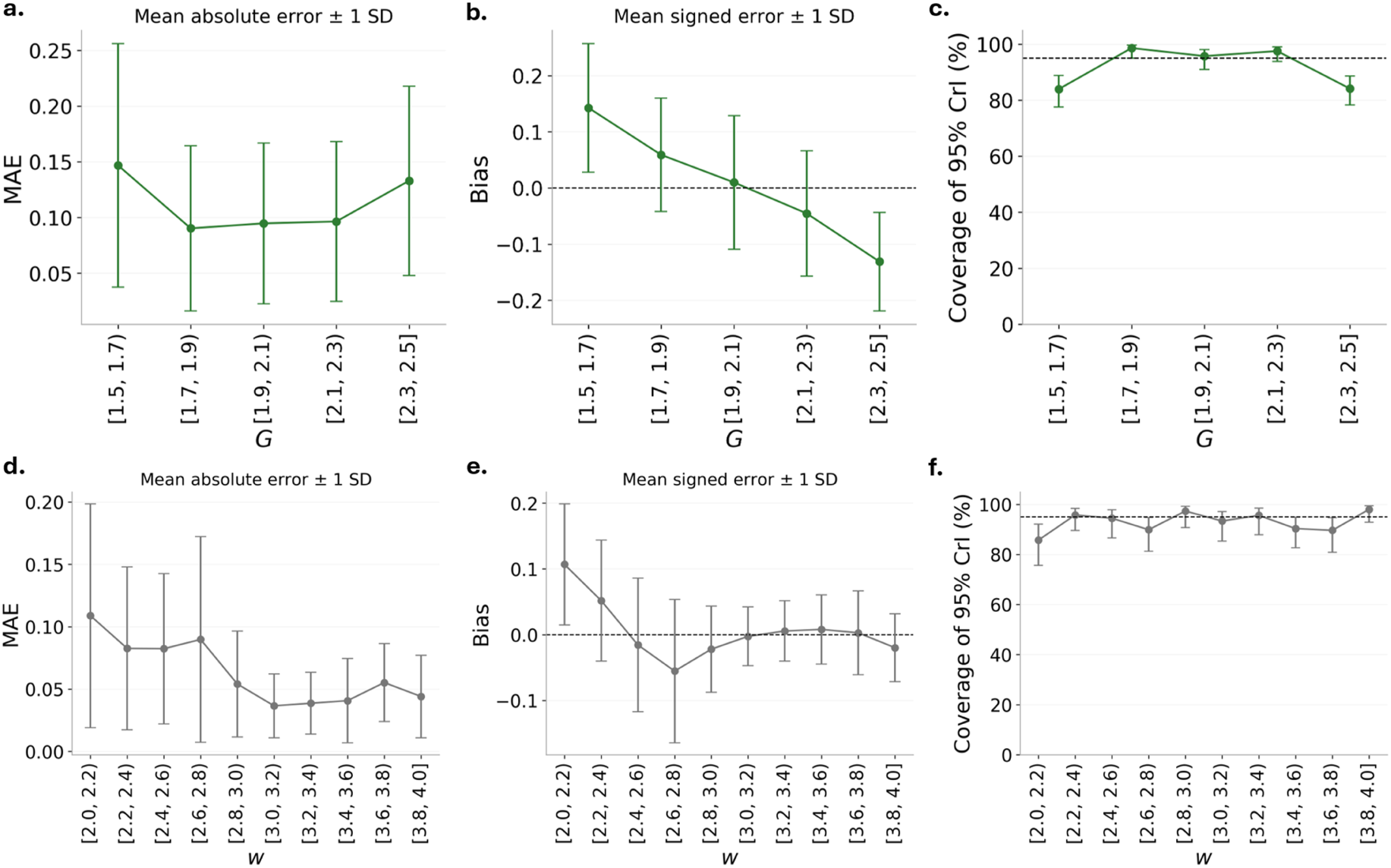
Parameter-dependent recovery accuracy and credible-interval coverage for the global inference model. Recovery diagnostics were evaluated across the held-out synthetic test set by stratifying simulations according to the ground-truth values of global coupling *G* (top row) and recurrent excitation *w* (bottom row). **(a,d)** Mean absolute estimation error within each parameter interval, with error bars indicating one standard deviation. **(b,e)** Mean signed estimation error, defined as the posterior mean minus the ground-truth value, with error bars indicating one standard deviation. The dashed horizontal line denotes zero bias. **(c,f)** Empirical coverage of the nominal 95% credible intervals within each parameter interval. The dashed horizontal line indicates the nominal 95% coverage level. The results reveal parameter-dependent variations in recovery accuracy and calibration, including increased estimation errors and reduced credible-interval coverage near portions of the prior boundaries.

**Figure SM4:**
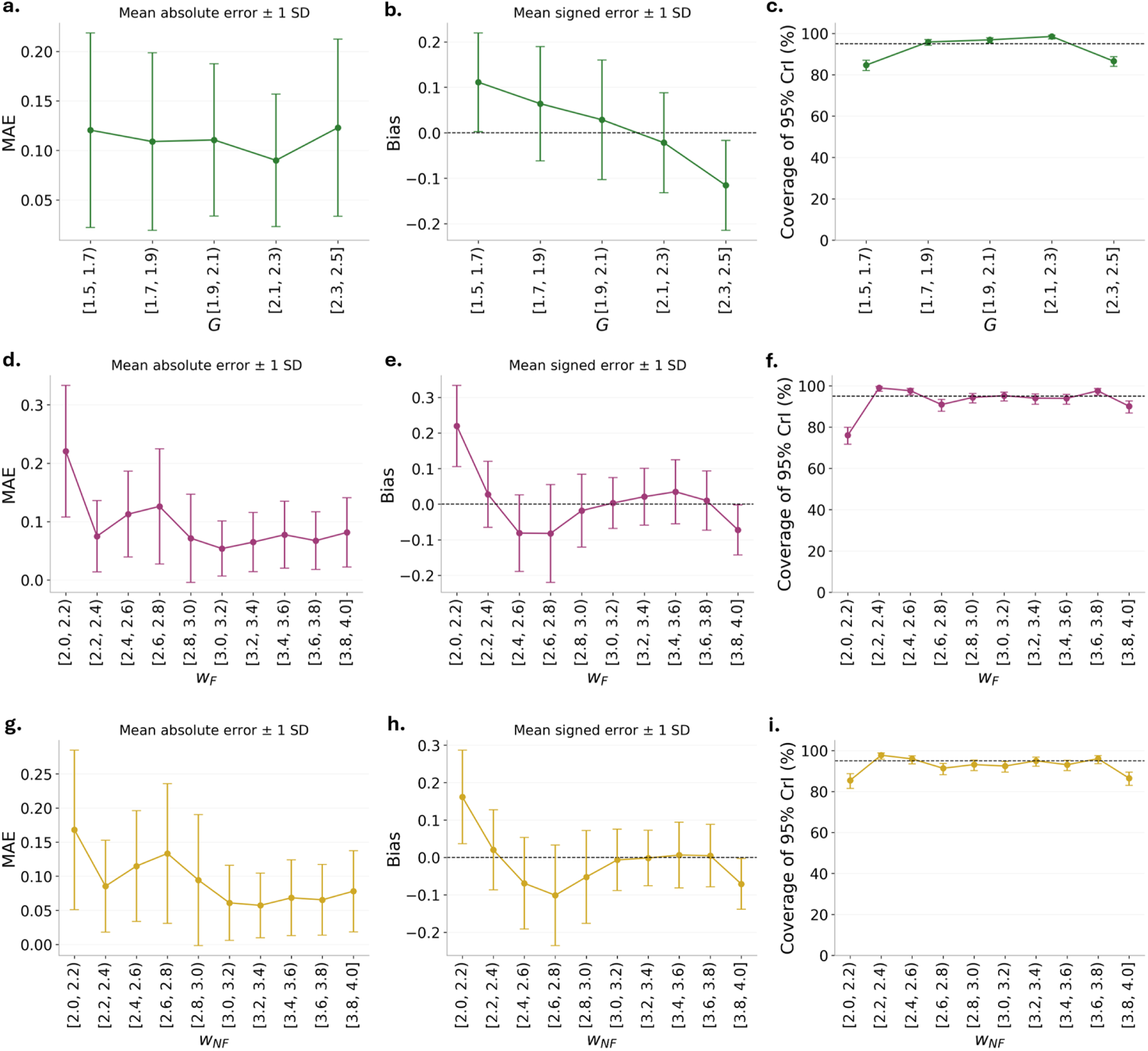
Parameter-dependent recovery accuracy and posterior coverage for the regional inference model. Recovery diagnostics were evaluated using 4,000 independent held-out synthetic simulations by stratifying the results according to the ground-truth values of global coupling (G) (top row), motor-frontal recurrent excitation (*w_F_*) (middle row), and non-frontal recurrent excitation (*w_NF_*) (bottom row). **(a,d,g)** Mean absolute estimation error within each parameter interval, with error bars indicating one standard deviation. **(b,e,h)** Mean signed estimation error, defined as the difference between the posterior mean estimate and the ground-truth parameter value, with error bars indicating one standard deviation. The dashed horizontal line denotes zero bias. **(c,f,i)** Empirical coverage of the nominal 95% credible intervals within each parameter interval. The dashed horizontal line indicates the nominal coverage level. Recovery accuracy and posterior calibration vary across the investigated parameter ranges, with increased estimation errors, systematic local biases, and reduced credible-interval coverage near some prior boundaries.

**Figure SM5:**
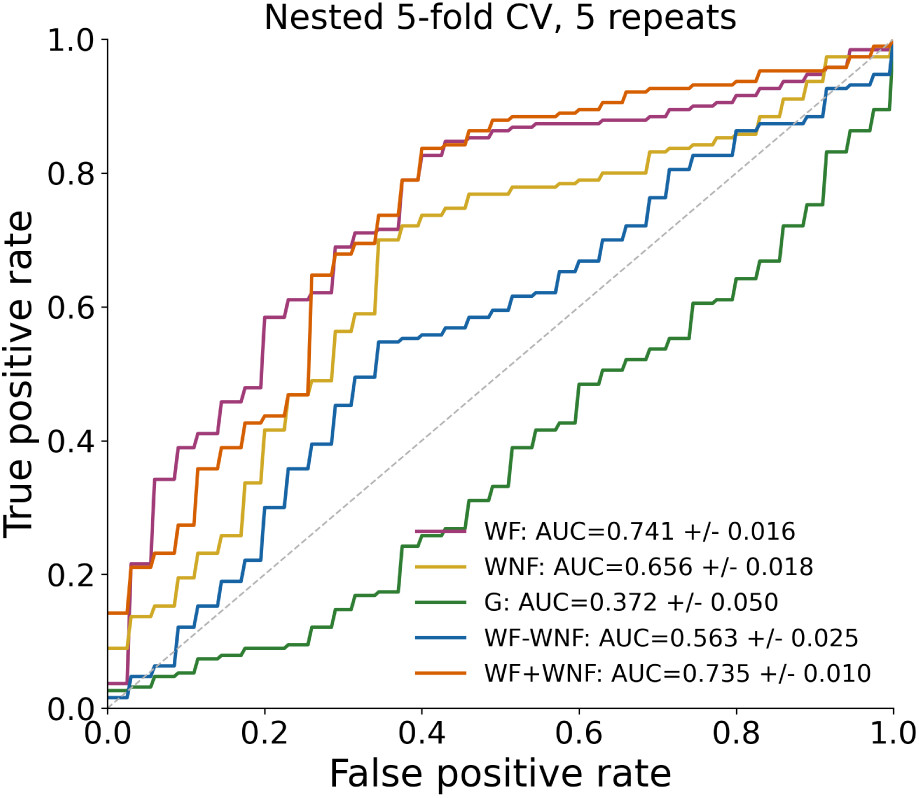
Classification performance of individual inferred parameters and their regional combinations. Receiver operating characteristic (ROC) curves for discriminating ALS participants (n = 38) from controls (n = 35), using individual posterior means of *G*, *w_F_* and *w_NF_* , or the derived quantities *w_F_* + *w_NF_* and *w_F_ −w_NF_* . Each predictor was evaluated separately using regularized logistic regression with nested stratified five-fold cross-validation repeated five times. Predictor standardization and hyperparameter selection were performed within the training folds, using identical outer splits across predictors. Legend values indicate the mean area under the ROC curve (AUC) standard deviation across repetitions. The dashed diagonal denotes chance-level discrimination. The regional sum yielded a numerically similar AUC to *w_F_* alone, whereas the regional difference showed lower discrimination.

**Figure SM6:**
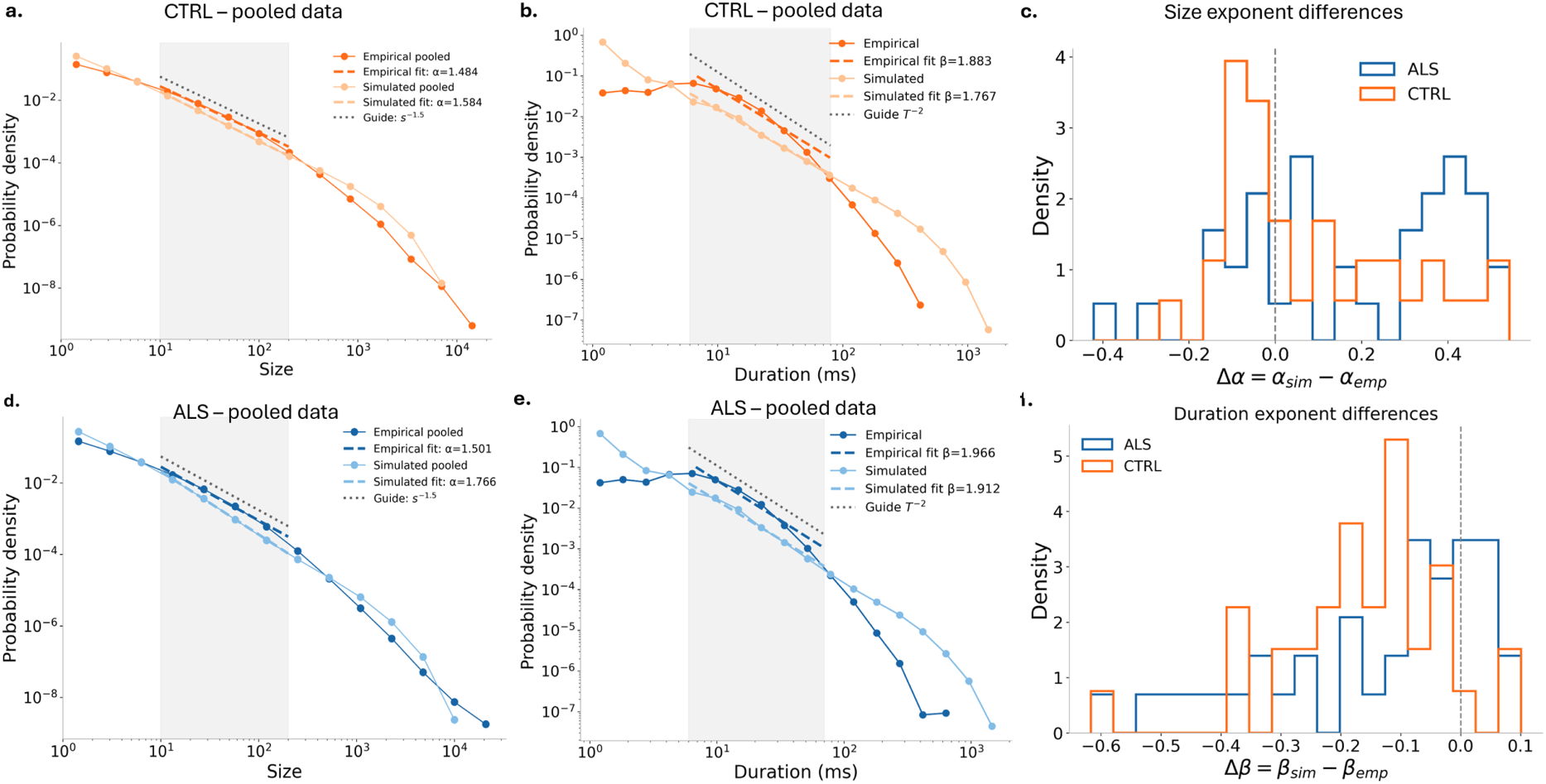
Comparison of empirical and simulated neuronal avalanche-size and duration distributions in ALS patients and healthy controls. One simulation was generated for each participant using the individual posterior mean estimates of motor-frontal recurrent excitation *w_F_* , remaining-region recurrent excitation *w_NF_* , and global coupling *G*. The analysis included 38 ALS patients and 35 controls with available inferred parameters. Avalanches were extracted independently from each recording and subsequently pooled within diagnostic groups. **(Left column:)** Pooled avalanche-size distributions for controls (**a**) and ALS patients (**d**). Avalanche size was defined as the cumulative amplitude excess above threshold, *S* = Σ*_r,t_* max(| *z_r_*(*t*)| 2.5, 0), over the samples and regions belonging to each avalanche. Curves include events with *S ≥*1 and are normalized over the retained events. Darker colors represent empirical data (orange for controls and blue for ALS), and lighter colors represent simulations. Dashed lines show continuous power-law fits, *P* (*S*) ∝ *S^−α^*, estimated from unbinned event sizes over 10 *≤S ≤*200 (shaded region). Gray dotted lines provide the visual reference *S^−^*^1.5^. **Middle column:** Corresponding pooled duration distributions for the CTRL group (**b**) and ALS patients (**e**) . Durations are expressed in milliseconds, using the native sampling intervals of the empirical and simulated recordings (1024 and 1000 Hz, respectively). Dashed lines show discrete power-law fits, *P* (*T*) ∝ *T ^−β^*; shaded regions indicate these fitting intervals. Gray dotted lines indicate the reference *T ^−^*^2^.**Right column:** Distributions of subject-level exponent differences for sizes (**c**), Δ*α* = *α*_sim_ − *α*_emp_, and durations (**f**), Δ*β* = *β*_sim_ *β*_emp_. Blue and orange outlines denote ALS and control participants, respectively. Vertical dashed lines mark zero difference. Positive differences indicate a steeper simulated decay, whereas negative differences indicate a slower simulated decay. Reference slopes and fitted exponents describe scaling within the selected intervals.

